# Transcranial Blood–Brain Barrier Opening in Alzheimer’s Disease Patients Using a Portable Focused Ultrasound System with Real-Time 2-D Cavitation Mapping

**DOI:** 10.1101/2023.12.21.23300222

**Authors:** Sua Bae, Keyu Liu, Antonios N. Pouliopoulos, Robin Ji, Sergio Jiménez-Gambín, Omid Yousefian, Alina R. Kline-Schoder, Alec J. Batts, Fotios N. Tsitsos, Danae Kokossis, Akiva Mintz, Lawrence S. Honig, Elisa E. Konofagou

## Abstract

**Background:** Focused ultrasound (FUS) in combination with microbubbles has recently shown great promise in facilitating blood-brain barrier (BBB) opening for drug delivery and immunotherapy in Alzheimer’s disease (AD). However, it is currently limited to systems integrated within the MRI suites or requiring post-surgical implants, thus restricting its widespread clinical adoption. In this pilot study, we investigate the clinical safety and feasibility of a portable, non-invasive neuronavigation-guided FUS (NgFUS) system with integrated real-time 2-D microbubble cavitation mapping.

**Methods:** A phase 1 clinical study with mild to moderate AD patients (N=6) underwent a single session of microbubble-mediated NgFUS to induce transient BBB opening (BBBO). Microbubble activity under FUS was monitored with real-time 2-D cavitation maps and dosing to ensure the efficacy and safety of the NgFUS treatment. Post-operative MRI was used for BBB opening and closure confirmation as well as safety assessment. Changes in AD biomarker levels in both blood serum and extracellular vesicles (EVs) were evaluated, while changes in amyloid-beta (Aβ) load in the brain were assessed through ^18^F-Florbetapir PET.

**Results:** BBBO was achieved in 5 out of 6 subjects with an average volume of 983±626 mm^3^ following FUS at the right frontal lobe both in white and gray matter regions. The outpatient treatment was completed within 34.8±10.7 min. Cavitation dose significantly correlated with the BBBO volume (*R*^2^>0.9, *N*=4), demonstrating the portable NgFUS system’s capability of predicting opening volumes. The cavitation maps co-localized closely with the BBBO location, representing the first report of real-time transcranial 2-D cavitation mapping in the human brain. Larger opening volumes correlated with increased levels of AD biomarkers, including Aβ42 (*R*^2^=0.74), Tau (*R*^2^=0.95), and P-Tau181 (*R*^2^=0.86), assayed in serum-derived EVs sampled 3 days after FUS (*N*=5). From PET scans, subjects showed a lower Aβ load increase in the treated frontal lobe region compared to the contralateral region. Reduction in asymmetry standardized uptake value ratios (SUVR) correlated with the cavitation dose (*R*^2^>0.9, *N*=3). Clinical changes in the mini-mental state examination over 6 months were within the expected range of cognitive decline with no additional changes observed as a result of FUS.

**Conclusion:** We showed the safety and feasibility of this cost-effective and time-efficient portable NgFUS treatment for BBBO in AD patients with the first demonstration of real-time 2-D cavitation mapping. The cavitation dose correlated with BBBO volume, a slowed increase in pathology, and serum detection of AD proteins. Our study highlights the potential for accessible FUS treatment in AD, with or without drug delivery.

## Introduction

Alzheimer’s disease (AD) is the most common neurodegenerative disorder, typically with progressive amnestic cognitive impairment, and its prevalence increases with the aged population growth [1]. Only since 2021 have effective disease-modifying treatments been available for Alzheimer’s disease. Currently, three monoclonal antibodies against amyloid-beta (Aβ) have been shown to be effective: aducanumab [2], lecanemab [3], and donanemab [4]. Clinical studies have demonstrated that administration of these antibodies markedly reduces the amount of Aβ-containing cerebral neuritic plaques, and the latter two antibodies have been proven to slow the clinical decline. However, these antibodies do have limitations, including only limited slowing of cognitive decline and cerebral side effects, such as amyloid-related imaging abnormalities of edema/effusion and hemorrhage. Anti-Aβ monoclonal antibodies bind to different forms of Aβ and promote clearance of plaques from the brain. However, for most, but not all, antibodies, the blood-brain barrier (BBB) poses a challenge to their use because only a small proportion of the administered drug is able to enter the brain, which affects their effectiveness in terms of dosage, frequency, and duration of treatment [5,6]. Although direct intracerebral infusion could possibly circumvent the BBB restriction, this invasive procedure entails risks [7].

Transient BBB opening by microbubble-mediated focused ultrasound (FUS) is a promising non-invasive therapy for enhancing BBB permeability, thereby facilitating the delivery of therapeutic drugs or promoting immune responses without the use of drugs [8–10]. In this treatment, microbubbles are systemically administered while FUS induces the rapid oscillation of microbubbles, called cavitation, at a targeted volume in the brain (Figure 1). Precise FUS treatment can induce local and transient BBB opening (BBBO) and promote immune response [11,12]. Numerous preclinical studies have proven that BBBO can lead to a decrease in Aβ or tau proteins in the brain and the cognitive improvement with and without drugs by increased immune response such as microglial phagocytosis [13–15].

**Figure 1.**
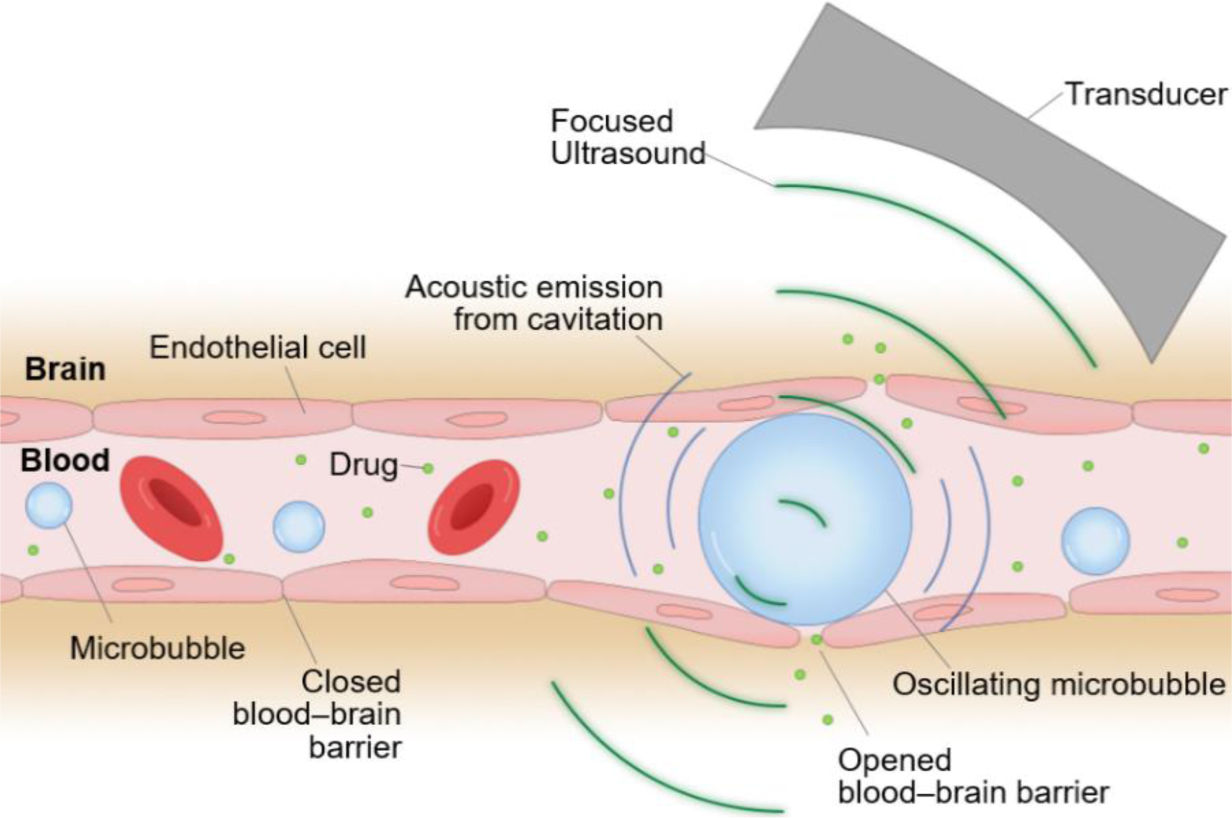
Illustration of focused ultrasound (FUS)-induced blood-brain barrier (BBB) opening. Systemically administered microbubbles oscillate under localized FUS and transiently open the BBB for drug delivery or immune-stimulation at the targeted brain tissue. Oscillating microbubbles emit acoustic cavitation signals which can be used for treatment monitoring.

Many clinical trials have demonstrated that magnetic resonance (MR)-guided focused ultrasound (MRgFUS) can safely and transiently open BBB in patients with AD [16–19], amyotrophic lateral sclerosis [20], Parkinson’s disease [21], glioma [22,23], and brain metastases [24]. Previous studies with AD patients have shown that BBBO induces a modest reduction in ^18^F-Florbetaben uptake ratio in PET with no cognitive worsening after multiple sessions of MRgFUS treatment [16–19,25]. A recent study utilizing MRgFUS reported that monthly BBBO treatment combined with aducanumab infusion led to a substantial reduction in Aβ levels in three patients over six months, as measured by PET [26]. While MRgFUS is the most widely used approach for clinical trials of BBBO, it generally requires the patient to stay still with their heads fixed by a stereotaxic frame in the MR scanner for a few hours. Alternatively, an implantable FUS device has been utilized in clinical trials, demonstrating a non-significant reduction in amyloid based on ^18^F-Florbetapir PET scans after multiple treatments [27]. Although this approach has been proven well-tolerated, it requires a burr hole achieved with brain surgery, making it invasive. The invasiveness of the procedure makes it a less preferred option for treating Alzheimer’s disease, particularly given that the typical patient population is older and may have multiple comorbidities.

Given the need for repetitive treatments and the advanced age of Alzheimer’s disease patients, there is a compelling demand for facilitating a low-cost and non-invasive treatment approach. Portable neuronavigation-guided FUS (NgFUS) systems can provide FUS treatment outside an MR scanner in an outpatient room. Portable systems have been employed in both preclinical and clinical studies [28–31], but only one clinical study has been reported in the context of Alzheimer’s disease, showing modest cognitive improvement after FUS [28]. However, this study did not induce BBBO or investigate if there were any changes in amyloid or tau protein load.

In this Phase 1 clinical study (NCT04118764), we assessed the clinical feasibility and safety of BBBO in six subjects with mild-to-moderate Alzheimer’s disease using a portable FUS system that we developed and verified in preclinical studies [32–34]. A single 2-minute FUS sonication session was performed per subject without a stereotaxic frame or a MR scanner. Real-time cavitation monitoring was employed to measure the treatment dose and assess its capability of predicting BBBO volume in the human brain. To our knowledge, this is the first report of real-time cavitation mapping in the human brain using a portable FUS system. Neurological and biological effects of the portable FUS system were evaluated by blood biomarker analysis, ^18^F-Florbetapir PET scans, and mini-mental state examination (MMSE).

## Results

### Study Overview

The primary objective of the study is to assess the safety and feasibility of FUS-induced BBBO in Alzheimer’s disease patients, using a portable, noninvasive FUS system. Secondary objectives includes testing the feasibility of cavitation mapping and observing changes in ^18^F-Florbetapir PET and in blood biomarkers. Six Alzheimer’s disease patients (2M/4F, age = 69.7±7.2 yr) were enrolled in a phase 1 trial under FDA and Columbia University IRB approval (NCT04118764) (Table 1). Inclusion and exclusion criteria, including diagnosis of Alzheimer’s disease, amyloid positivity on ^18^F-Florbetapir PET scan, and an MMSE score between 12 and 26, are listed in Table S1. Figure 2 presents the timeline of the clinical trial. All subjects underwent one session of FUS sonication and had pre- and post-treatment scans, blood tests, and MMSE assessments.

**Figure 2.**
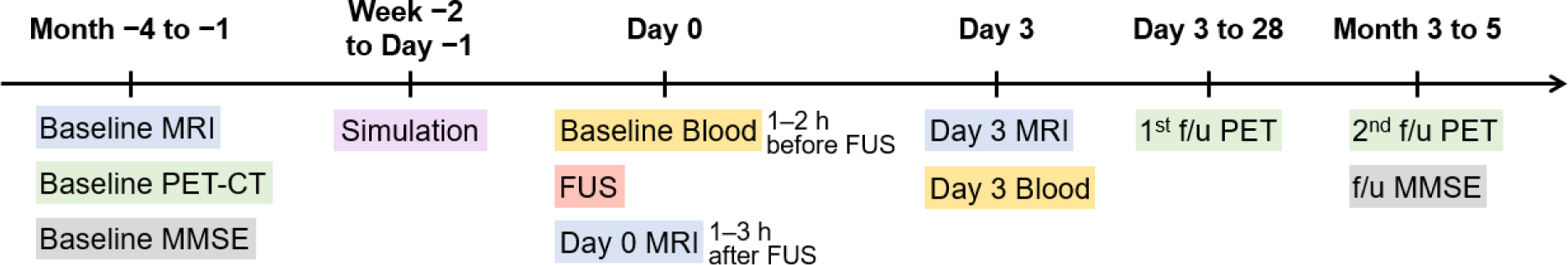
Timeline of the clinical study.

**Table 1.** Patient Characteristics.

| Subject# | 1 | 2 | 3 | 4 | 5 | 6 |
| --- | --- | --- | --- | --- | --- | --- |
| Age range | 70-74 | 80-84 | 65-69 | 65-69 | 55-59 | 70-74 |
| Sex | F | M | F | F | M | F |
| MMSE | 18 | 20 | 15 | 21 | 24 | 15 |
| GDS | 1 | 1 | 2 | 1 | 0 | 0 |
| MHIS | 1 | 0 | 2 | 1 | 0 | 1 |

MMSE: Mini-Mental State Examination on the day of screening, GDS: Geriatric Depression Scale on the day of screening, MHIS: Modified Hachinski Ischemia Scale

### Portable NgFUS system allowed for efficient BBBO

The target location was selected in a PET amyloid-positive region of the right frontal lobe, and the FUS trajectory was determined by considering the beam incidence angle relative to the skull (Figure 3A). Skull insertion loss (i.e., attenuation), obtained from patient-specific acoustic simulations (Figure 3B) and shown in Table 2, allowed us to adjust the sonication power to deliver a derated in situ peak-negative pressure of 200 kPa. On the day of treatment, all subjects received a single FUS treatment with microbubble administration (0.1 mL/kg) using the portable NgFUS system, while seated in a medical recliner chair in an outpatient unit (Figure 3C). This process was guided by neuronavigation (Figure 3D) and monitored by cavitation dose and mapping (Figure 3E). FUS was deployed through a contact area with a diameter of less than 50 mm, which allowed for partial hair shaving instead of complete head shaving (Figure S1). All sessions were uneventful and the average treatment procedure time was 34.8±10.7 min.

**Figure 3.**
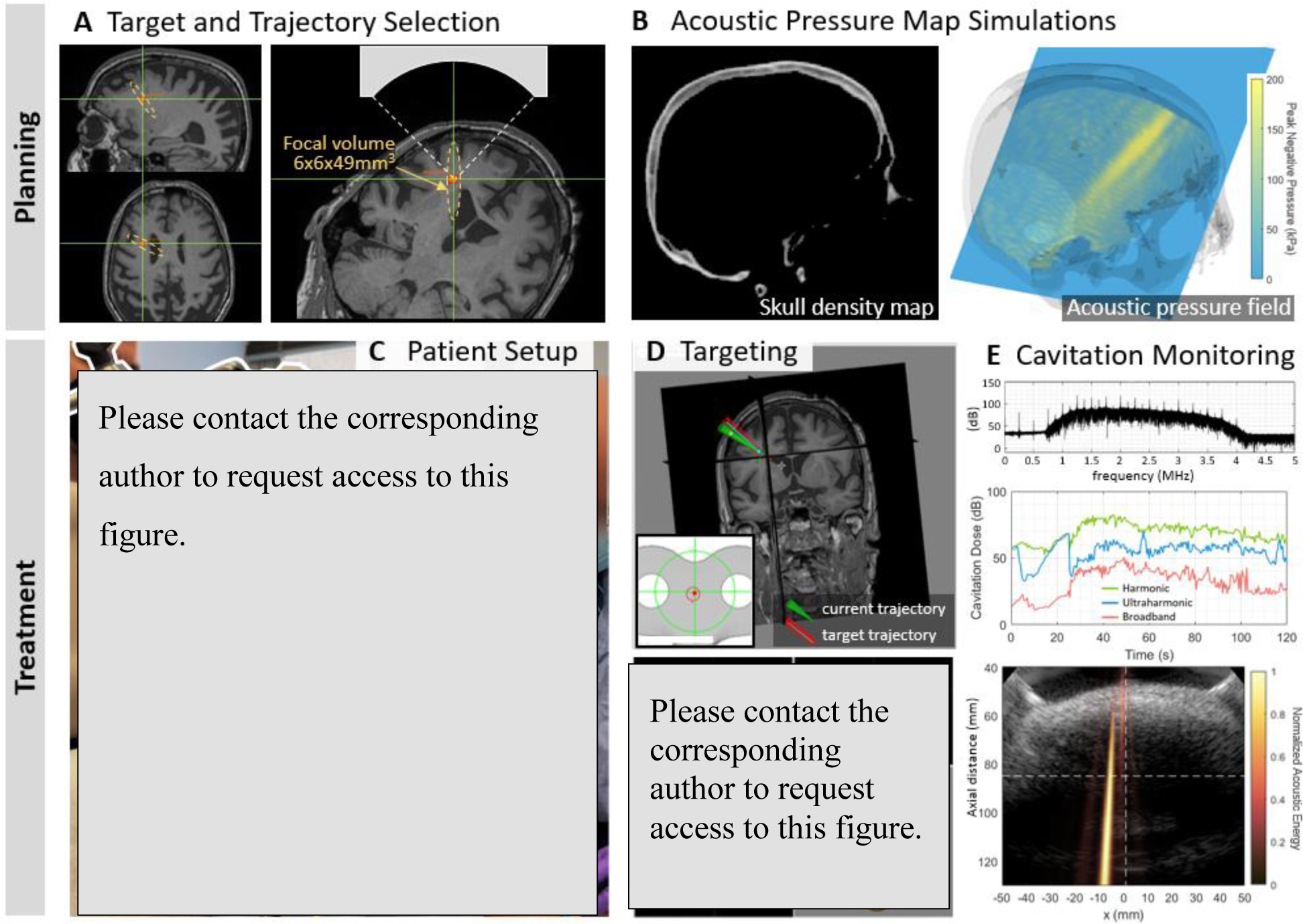
Portable NgFUS planning and treatment. **(A)** Selection of the target and the trajectory of the FUS beam. **(B)** Acoustic pressure map (right) obtained from simulations using CT image (left) for skull insertion loss estimation. **(C)** Subject undergoing the treatment session. The subject’s head was supported and fixed by a head and chin rest. The FUS transducer with a coaxial single-element transducer (N=4) or a phased array transducer (N=2) was fixed with the metallic arm during the 2-min treatment. **(D)** Targeting with the real-time feedback of the neuronavigator. **(E)** Real-time cavitation monitoring with the frequency spectrum, cavitation dose, and cavitation energy map (color) with the B-mode image (grayscale) (from top to bottom). With PCD monitoring, only the frequency spectrum and cavitation dose were obtained. With PAM, a cavitation map was obtained as well as the spectrum and dose.

**Table 2.** Summary of the treatment.

| Subject # | 1 | 2 | 3 | 4 | 5 | 6 |
| --- | --- | --- | --- | --- | --- | --- |
| Contrast-enhanced volume (mm<sup>3</sup>) | 2013 | 414 | 0 | 951 | 278 | 1262 |
| Skull-induced attenuation* | 0.84 | 0.72 | 0.75 | 0.72 | 0.7 | 0.75 |
| Procedure time** (min) | - | 38 | 47 | 21 | 36 | 35 |
| Transducer positioning errors† (mm, °) | - | 4.7, 11.2 | 5.3, 13.0 | 4.3, 7.0 | 8.2, 14.5 | 5.8, 10.5 |
| Subject movements†† (MAD, max) (mm) | - | - | 2.3, 9.4 | 0.5, 1.5 | 0.25, 1.1 | 0.28, 0.96 |
| Distance between the BBBO centroid and the simulated focus‡ (mm) | 23.2 | 13.3 | - | 8.4 | 5.4 | 11.3 |

\* Obtained by the acoustic simulation based on the planned target prior to the treatment. \*\* Time duration for the subject sitting on the treatment chair; including anatomical registration for the neuronavigation, targeting, sonication, and patient release. † Distance (mm) and angle difference (°) between the planned trajectory and the actual trajectory during the treatment. †† Subject movement was measured by the mean absolute deviation (MAD) from its centroid and the maximum distance (max) from the initial location. ‡ Distance between the centroid of the BBBO and the estimated focal position in the simulated pressure map

Five subjects underwent successful BBBO at the treated location in the frontal lobe as evidenced by post-FUS T1-weighted MRI (Figure 4A), and the quantified contrast-enhanced volume, which serves as a measure of BBBO volume, was 983±626 mm^3^ (Figure 4C and 4D and Table 2). One participant (subject 3) had no detectable opening (Figure S2), likely due to inadequate microbubble administration caused by a syringe malfunction and patient movement resulting from not using the head and chin rest. Subject 1 exhibited the largest opening volume of 2,013 mm^3^, which extended to the left thalamus beyond the lateral ventricles while subject 4 exhibited the smallest opening volume of 278 mm^3^. All of the openings from the 5 subjects were closed within 72 h, which was confirmed by the follow-up scans on day 3 (Figure 4B). The opening and closing of the BBB were confirmed by a neuro-radiologist, and the BBBO was quantified from the subtracted contrast-enhanced T1-weighted MRI, as described in the Materials and Methods section.

**Figure 4.**
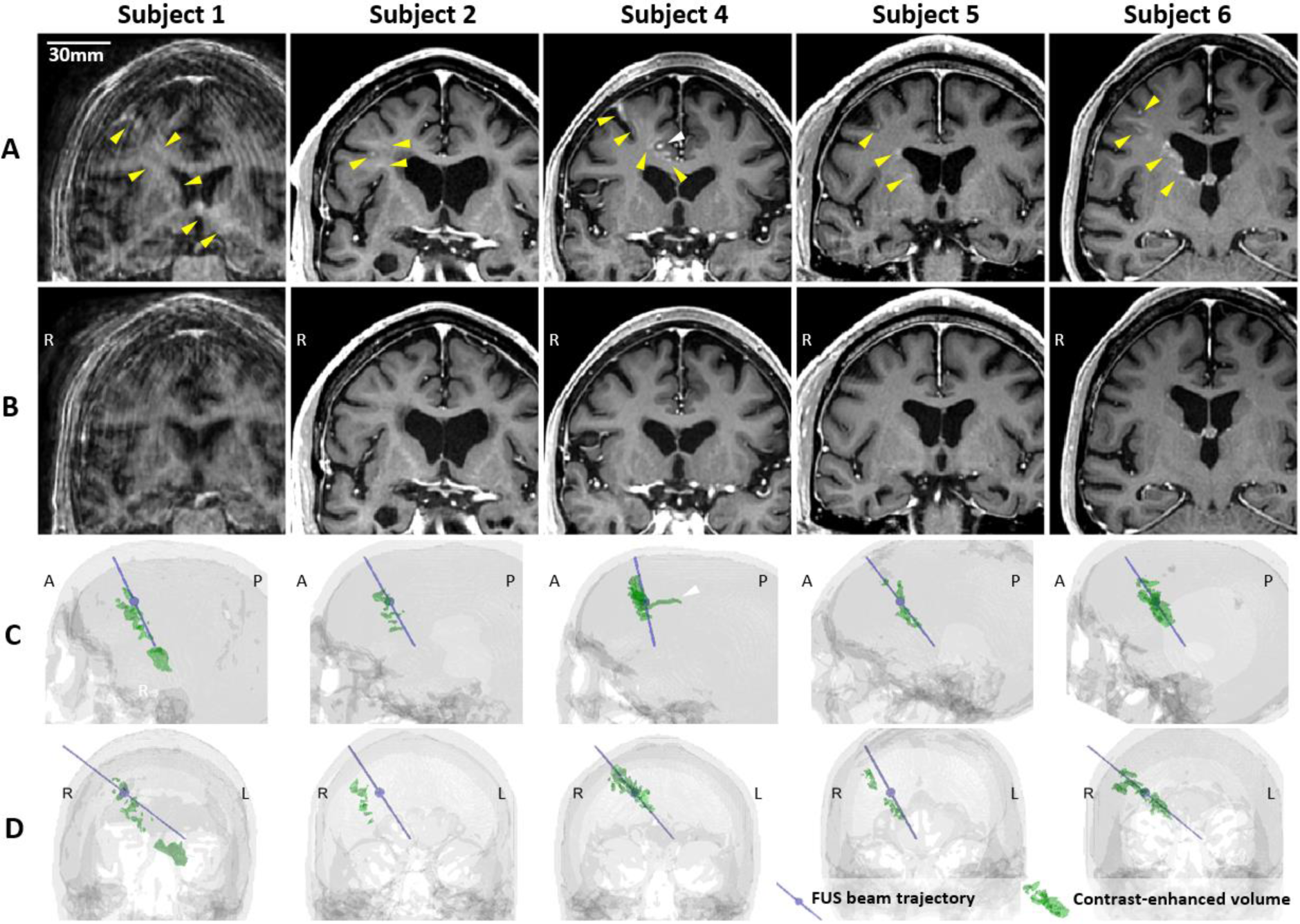
Blood-brain barrier (BBB) opening and closure confirmed by contrast-enhanced T1-weighted MRI. (A) Contrast enhancement indicating BBB opening in T_1_-weighted MRI 2 hours after focused ultrasound (FUS) sonication. (B) Lack of contrast enhancement detected on follow-up T_1_-weighted MRI confirmed BBB reinstatement on day 3. (C, D) The 3-dimensional (3-D) reconstruction of the FUS beam trajectory (blue line) and segmented contrast-enhanced volume (green) overlaid on the CT skull image (gray) in (C) the sagittal and (D) the coronal view. The maximum pressure point of the focus is denoted as a blue sphere on the blue line. The contrast-enhanced volume was well aligned with the FUS beam trajectory for subjects 4–6. For subjects 1 and 2, the opening was aligned with the trajectory in the sagittal plane (C) but approximately 10 mm off from the trajectory in the coronal plane (D). A video for 360° view of 3-D volumes is available online as Supplementary Movie 1.

### Safety evaluation

There were no serious adverse events (SAEs) and no clinical changes after the treatments. One subject (subject 1) had an adverse event (AE) including both mild skin erythema on day 0 (resolved within 3 days) and asymptomatic cerebral edema with a superficial hemorrhagic component on day 3. MRI images showed an area of T2 fluid-attenuated inversion recovery (FLAIR) hyper-intensity on day 3, most intense at the cortical targeted location but extending deeper (Figure S3), with susceptibility-weighted imaging (SWI) hypo-intensity superficially within the same region (Figure S4). The subject was asymptomatic and the MRI abnormalities were all resolved in follow-up scans on day 15. Other subjects did not show abnormalities in the safety MR scans 3 days after the treatment.

### Cavitation dose and map showed promising results for predicting the BBBO

To monitor the safety and efficacy of the treatment, cavitation signal during sonication was observed. Passive cavitation detection (PCD) with a single-element transducer was utilized to obtain cavitation dose (CD) for subjects 1–4, while passive acoustic mapping (PAM) with a imaging array transducer was employed for subjects 5 and 6 to obtain the 2-D cavitation maps.

Figures 5A–5C show the real-time cavitation dose monitoring results from subjects 1–4. For subjects 1, 2, and 4, ultraharmonic and broadband CDs increased after the microbubble injection (*t* = 20–30 s) and persisted until the end of the sonication (*t* = 120 s), indicating the cavitation activity of the injected microbubbles (Figure 5A). On the other hand, for subject 3 which exhibited no detectable opening, the increased CD was not sustained over time, resulting in a low cumulative CD (CCD). The low cavitation energy for subject 3 is also evident in the spectrogram (Figure 5B). Across the four subjects, the higher CCDs were detected with increasing BBBO volume (Figure 5C), resulting in strong positive linear correlations (*R*^2^ > 0.9, *p* < 0.05).

**Figure 5.**
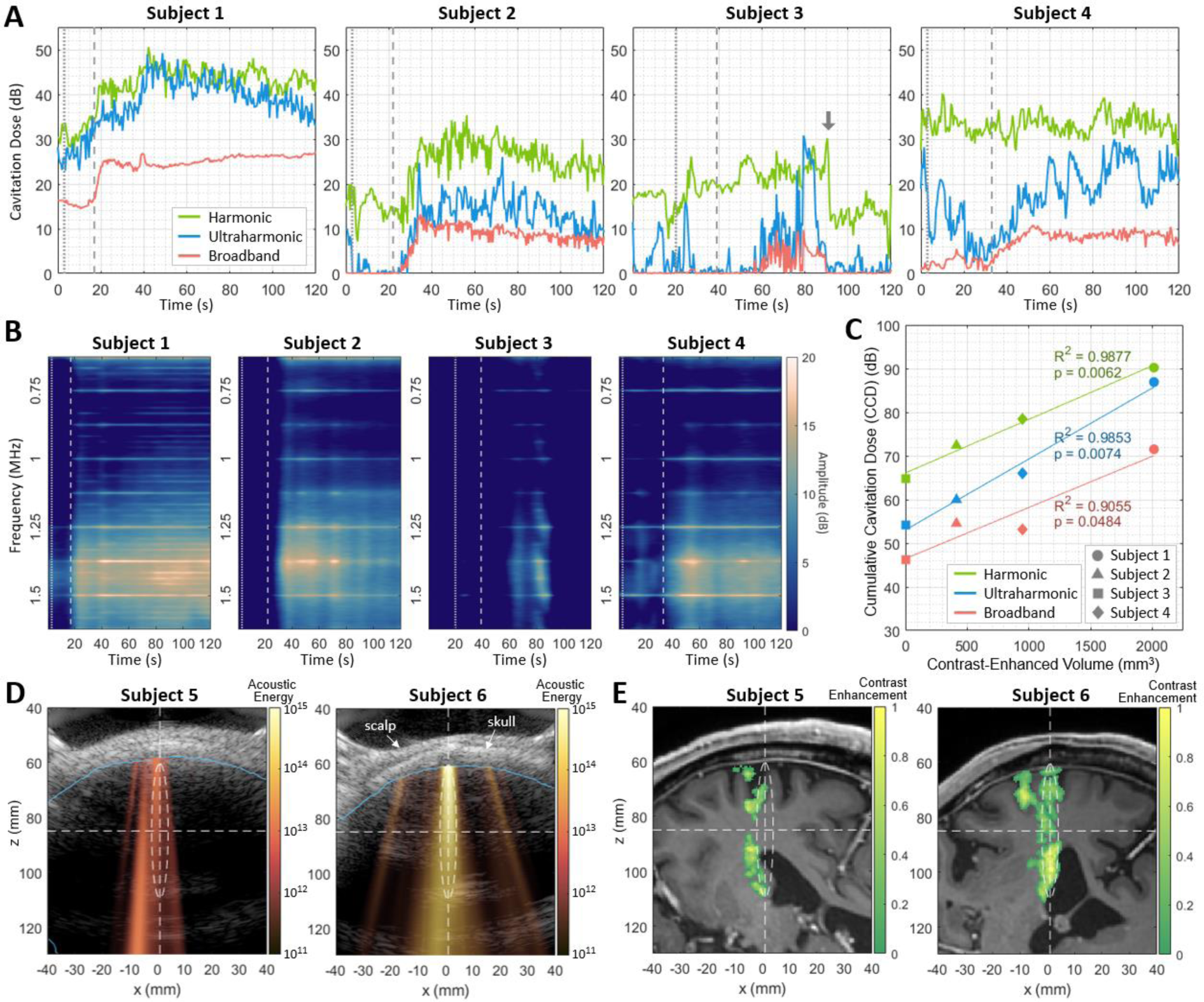
Real-time cavitation monitoring during focused ultrasound (FUS) treatment. **(A)** Harmonic, ultraharmonic, broadband cavitation doses (CDs) during the sonication. The CDs of subjects 1, 2, and 4 increased after the microbubble injection and flush and were sustained until the end of sonication. In contrast, the CDs of subject 3, who did not exhibit successful BBBO, were unstable and exhibited a sudden reduction at t = 90 s. The gray arrow indicates the moment of a sudden subject movement detected (Supplementary Figure S7A). **(B)** Spectrograms displayed during the sonication showed the increased cavitation signal in subjects 1, 2, and 4. Vertical dotted and dashed lines in (A) and (B) indicate the time of the microbubble bolus injection and the subsequent saline flush, respectively. The amplitude in (B) was normalized by the baseline broadband cavitation dose to better represent harmonic and ultraharmonic components. **(C)** Positive correlation between the BBBO volume (i.e., contrast-enhanced volume) and the cumulative CDs (CCDs) over time. **(D)** Cavitation map (color), which presents the distribution of acoustic cavitation energy, is overlaid on the corresponding ultrasound B-mode image (gray) that shows the scalp and the skull profiles. The brain region obtained from the registered MRI is marked as a blue line. **(E)** Projected contrast-enhanced volume (color) overlaid on the MRI slice that is registered to the cavitation map/B-mode image in (D). White dashed lines and ellipsoids in (D) and (E) show the focus of the FUS beam.

Figures 5D and 5E depict the real-time 2-D cavitation maps obtained by PAM and their corresponding BBBO regions in MRI, respectively, from subjects 5 and 6. To our knowledge, this is the first demonstration of 2-D transcranial PAM in the human brain. The cavitation map that shows the spatial distribution of acoustic energy detected from microbubble activity (Figure 5D) roughly matched with the BBBO location (Figure 5E); both the acoustic energy and BBBO locations were shifted to the left side of the focus in subject 5, and aligned with the focus in subject 6. Compared to subject 5, subject 6 showed approximately 12 dB higher averaged acoustic energy in the map and exhibited a larger opening (278 mm^3^ vs. 1262 mm^3^). From the pixel-wise correlation analysis between the PAM cavitation map and the BBBO volume observed in MRI, the area under the curve (AUC) of the receiver operating characteristic (ROC) and precision-recall (PR) curves were AUC_ROC_=0.8 and AUC_PR_=0.7, respectively, showing the potential of PAM for predicting BBBO volume.

### Elevated blood biomarker levels correlated with BBBO size

Both serum and serum-derived extracellular vesicle (EV) levels of biomarkers 3 days after FUS were compared with the baseline levels obtained 1–2 hours prior to NgFUS for subjects 2–6. Subject 1’s biomarker levels were not obtained due to improper handling of the blood specimen and were excluded from the analysis. Figure 6 shows the correlation between BBBO volume and biomarker levels for S100β in serum, and Aβ42, Aβ42/Aβ40, GFAP, Tau, and pT181 in EVs. Subjects with larger opening volumes displayed elevated serum levels of S100 calcium-binding protein β (S100β) (*p* < 0.05), indicating compromised BBB integrity [35] (Figure 6A). Furthermore, we identified several statistically-significant positive linear relationships between the opening size and the serum-derived EV levels of Alzheimer’s disease-related proteins, while such relationships were not observed with serum biomarker levels. Notably, glial fibrillary acidic protein (GFAP), Tau, and phosphorylated-Tau 181 (pT181) fold-changes exhibited significant linear correlations (*p* < 0.05) (Figure 6D−6F), while the correlations for Aβ42 (*p* = 0.062) and the Aβ42/Aβ40 ratio (*p* = 0.096) were not statistically significant (Figure 6B and 6C). There were no significant group-wise changes possibly due to the large variation in BBBO volume (Figure S5).

**Figure 6.**
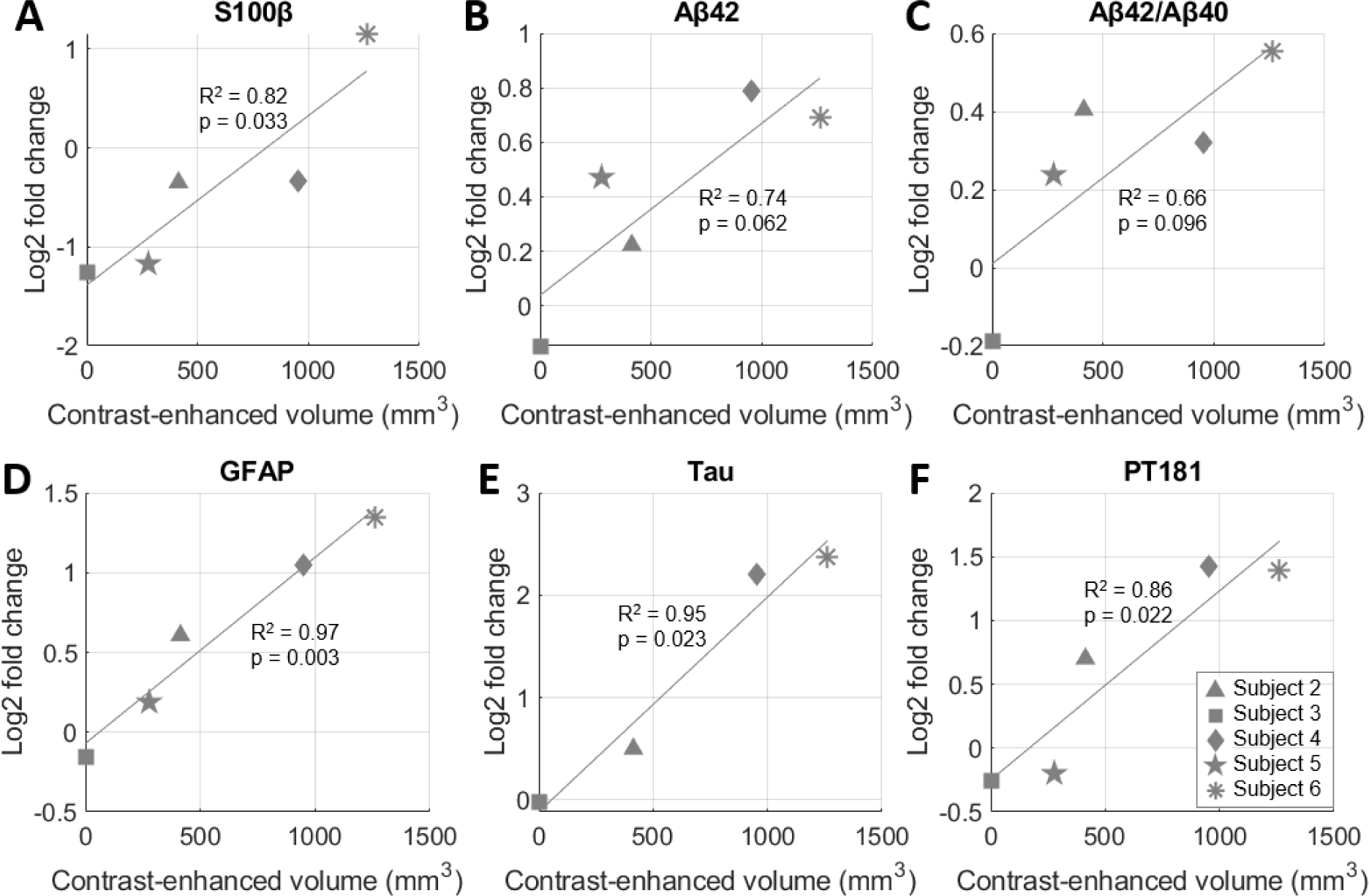
Correlation of blood-brain barrier opening (BBBO) volume and blood biomarker levels on day 3 after neuronavigation-guided FUS (NgFUS). A larger BBBO volume is associated with an increased log-fold change in biomarker concentration three days after treatment for **(A)** S100β in serum, **(B)** Aβ42, **(C)** Aβ42/Aβ40, **(D)** GFAP, **(E)** Tau, and, **(F)** pT181 in extracellular vesicles.

### A modest decrease in asymmetry SUVR correlated with the size of BBBO and cavitation dose

Figure 7 shows the percent changes in standard uptake value ratio (SUVR) or asymmetry SUVR of ^18^F-Florbetapir compared to the baseline. Subject 3 was excluded due to the absence of BBBO. Although there was no group-wise reduction in SUVR (Figure 7A–7C, Table 3), all subjects with BBBO showed a modest reduction in asymmetry SUVR which assesses the SUVR in the treated region compared to that of the contralateral region (Figure 7D–7F, Table 3). Specifically, asymmetry values decreased by 1.47±0.77% (*p*=0.013) in the frontal lobe and by 0.90±0.26% (*p*=0.001) in the hemisphere at the 2nd follow-up compared to the baseline. A non-significant linear relationship (*R*^2^=0.69, *p*=0.08) was measured between the BBBO volume and the 1st follow-up asymmetry changes within the respective volumes (Figure 7G). A relationship of the asymmetry SUVR change with CD was analyzed among subjects who exhibited BBBO with cavitation monitoring using a single-element detector (subjects 1, 2 and 4). Although negative linear relationships were observed between the 1st follow-up asymmetry changes and the CCDs (Figure 7H), these results should be interpreted with caution due to the small sample size (Figure 7H). Changes in SUVR and asymmetry values for each subject are listed in Table S2 and PET images are presented in Figure S2. SUVR Changes in Centiloid units are shown in Figure S6 to enable standardized comparison with other studies.

**Figure 7.**
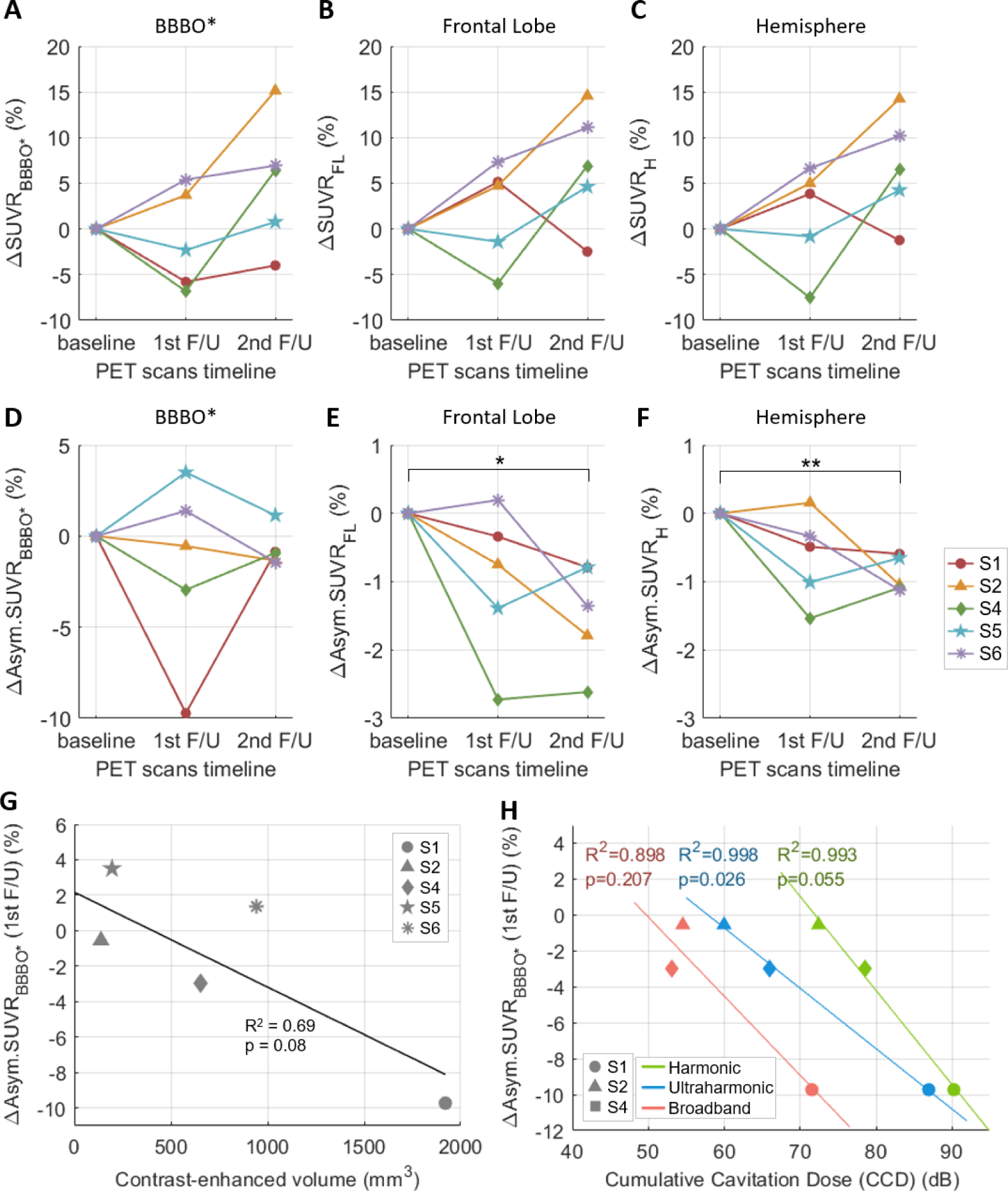
Percent changes in the standard uptake value ratio (SUVR) and asymmetry SUVR of ^18^F-Florbetapir and the correlation between the change in the asymmetry SUVR and the cumulative cavitation dose (CCD). **(A–C)** Percent changes in SUVR within the blood-brain barrier opening (BBBO) volume in the gray and white matter (ΔSUVR_BBBO*_), the right frontal lobe (ΔSUVR_FL_), and the right hemisphere (ΔSUVR_H_), at the 1st and the 2nd follow-ups compared to the baseline. **(D–F)** Percent changes in asymmetry SUVR (Asym.SUVR) within the BBBO volume in the gray and white matter (ΔAsym.SUVR_BBBO*_), the right frontal lobe (ΔAsym.SUVR_FL_), and the right hemisphere (ΔAsym.SUVR_H_) compared to the baseline. Significant reduction in asymmetry values were found when measured within the (E) frontal lobe and (F) hemisphere regions. *p<0.05, **p<0.01. **(G)** Linear correlation between the ΔAsym.SUVR_BBBO*_ and the BBBO volume in the gray and white matter. **(H)** Linear correlations of ΔAsym.SUVR_BBBO*_ with harmonic, ultraharmonic, broadband CCDs.

**Table 3.** Changes in SUVR and asymmetry SUVR at the 1st and 2nd follow-up compared to the baseline (mean±std)

| Region | ΔSUVR | | ΔAsym.SUVR | |
| --- | --- | --- | --- | --- |
|  | 1st F/U | 2nd F/U | 1st F/U | 2nd F/U |
| BBBO* | -1.17±5.51, p=.66 | 5.05±7.20, p=.19 | -1.67±5.09, p=.50 | -0.68±1.05, p=.22 |
| FL | 1.97±5.51, p=.47 | 6.95±6.51, p=.076 | -1.00±1.12, p=.12 | -1.47±0.77, p=.013 |
| H | 1.43±5.73, p=.61 | 6.80±5.88, p=.061 | -0.64±0.65, p=.092 | -0.90±0.26, p=.001 |

SUVR: standard uptake value ratio, Asym.: asymmetry, BBBO\*: blood-brain barrier opening in gray and white matter, FL: frontal lobe, H: hemisphere, F/U: follow-up. The 1st F/U and 2nd F/Us were 20±9 days and 103±30 days after focused ultrasound treatment, respectively. Mean and standard deviations were calculated from 5 subjects with BBBO.

### A single BBBO treatment did not significantly alter cognitive function

Comparing the baseline scores, the MMSE score decreased by 1.80±2.71 among the five subjects with BBBO and by 2.50±2.93 among all six enrolled subjects, approximately 3 months after NgFUS. When compared with the Alzheimer’s disease neuroimaging initiative (ADNI) database, subjects with successful BBBO did not exhibit statistically different changes in MMSE over a similar time frame (*p* > 0.45), indicating no cognitive changes due to FUS-induced BBBO (Table S3). Individual MMSE scores are listed in Table S4.

### Targeting accuracy and precision

Targeting of the FUS transducer during treatment was performed using a manual arm. The distance and angular errors of manual transducer positioning were 5.7±1.4 mm and 11.2±2.5°, respectively. Mean absolute deviation and maximum distance of the subject motion during the 2-min treatment were 0.3±0.1 and 1.2±0.2 mm when using a head and chin rest (subjects 4–6), and 2.3 mm and 9.4 mm without using the rest (subject 3) (Figure S7, Table 2). Distance between the centroid of the BBBO and the simulated focus was 12.3±6.1 mm, mostly along the FUS trajectory. There was no consistent trend in the shift towards or away from the transducer, as it depended on the skull properties and the selected treatment location for each subject.

## Discussion

In this pilot study, we demonstrated the feasibility and safety of the portable FUS system with cavitation monitoring for BBB permeability enhancement at the right frontal lobe in six Alzheimer’s disease patients. Five out of six subjects underwent localized BBBO which resolved within 72 h. One AE occurred and resolved in 15 days, and no SAE was reported. No clinically significant changes were observed in SUVR or cognitive test scores after FUS; however, the asymmetry SUVR reduced modestly and exhibited a linear correlation with the BBBO volume. The CD demonstrated correlations with the BBBO volume, blood biomarker level increase, and the asymmetry SUVR decrease, proving the potential benefits of integrated cavitation monitoring to predict treatment outcomes.

In clinical studies using MRgFUS, acoustic cavitation dose maps were generated by sonicating dozens of subspots and mapping the measured cavitation dose to each subspot, assuming that the received cavitation signals originated only from the focus, neglecting off-site cavitation [23,24,36]. In contrast, our portable FUS system provides a real-time 2-D cavitation map, offering not only off-target cavitation imaging but also a more precise estimation of CD localized at the target. To our knowledge, this is the first demonstration of 2-D PAM cavitation maps being obtained in the human brain during BBBO. Cavitation mapping holds significant potential for enhancing both the safety and efficacy of future treatments. By providing real-time visualization of cavitation activity, this technology enables clinicians to immediately identify and mitigate any off-target effects, reducing the risk of unintended damage to surrounding tissues. Furthermore, improved accuracy in estimating the CD could result in more predictable and effective BBBO.

Although we aimed to deliver the same acoustic pressure at the brain target, the BBBO size varied among the subjects from 278 mm^3^ to 2013 mm^3^. This variability could be attributed to the challenges in estimation of skull-induced attenuation, which determined the FUS transmit power. The attenuation estimates may have been affected by CT-MR registration errors in the acoustic simulation (5–10 mm in distance and 1–16° in angle) that we identified at the conclusion of the trial. Another factor may lie in the transducer positioning errors which caused discrepancies between the trajectory used for the simulation and the achieved trajectory during treatment (Table 2). Further analysis of the pressure estimation errors is presented in the Supplementary Discussion. The re-estimated pressure map after correcting the errors and the trajectories showed higher acoustic energy for larger opening cases (Figure S8A), with the most intense energy distribution in subject 1. The re-estimated maximum pressure in the brain (Table S5) showed a linear relationship (*R*^2^ = 0.84) with the contrast-enhanced volume (Figure S8B), which may explain the variance in BBBO size across the subjects and the AE in subject 1.

Five subjects showed contrast enhancement within an ellipsoidal volume along the FUS beam trajectory (Figure 4C and 4D, and Movie S1) consistent with the cigar-shaped focus (Figure S8A). Although the FUS focal volume included more white matter (WM) than gray matter (GM), the opening volume exhibited similar proportions of GM and WM (Figure S9). The higher probability of opening in GM than in WM was reported in our previous non-human primate (NHP) studies [33,37,38], where GM exhibited increased susceptibility to BBBO relative to WM, indicated by increased contrast enhancement on T1-weighted MRI. In this study, notable sulcal enhancements in GM were observed (Figure 4A and Figures S10A and S10B). Additionally, the non-uniformly distributed opening within the focal volume may be explained by the regional difference in vascular density and tissue property, or increased ultrasound attenuation [37,39–42].

In subject 4, the contrast enhancement on MRI was found not only along the FUS trajectory from the superior frontal cortex to the cingulate cortex, but also along the cingulate sulcus in the anterior-posterior direction (indicated with white arrowheads in Figures 4A and 4C, and S9C). The sulcal enhancement beyond the ellipsoidal focus might not indicate BBBO, because it extended approximately 22 mm posterior from the focus while the focal size is only 6 mm wide. Instead, this vessel-like extravasation of the contrast agent might have occurred through the vessel wall or the perivascular space (PVS) that extends along the perforating vessels [43], indicating a potential increase in permeability of the blood-meningeal barrier. A possible explanation might be that the contrast agent entered the PVS through the disrupted BBB within the ellipsoidal focus and then permeated posteriorly along the cingulate sulcus. This finding may be consistent not only with recent preclinical studies on glymphatic clearance effect of microbubble-mediated FUS in rodents [44,45] but also with clinical studies using MRgFUS [46,47]. These clinical studies also demonstrated blood-meningeal barrier opening and glymphatic clearance in humans, reporting hyperintense linear enhancement along the hippocampal fissure [47] and contrast accumulation in the subarachnoid space at the frontal lobe [46] following BBBO.

The first possible reason for the BBBO failure in subject 3 is the delayed bolus injection of compromised microbubbles due to the malfunction of the syringe/catheter system. The catheter was blocked at the initial injection attempt, resulting in the pressurization inside the syringe and the destruction of the microbubbles. Although the microbubble solution was eventually injected at *t* = 20 s after the start of sonication, the increases in CDs were minimal compared to other subjects (Figures 5A and 5B). The second reason could be attributed to subject movement. The head and chin rest were not used for this subject, resulting in the medial movement of ∼9 mm during the 2-min sonication, which was approximately 6–10 times larger than those of other subjects (Figure S7 and Table 2). The subject movement (i.e., movement of the focus) might have compromised the localized acoustic energy delivered, as evidenced by the reduced CDs at *t* = 90 s coinciding with the sudden movement (gray arrows in both Figure 5A and Figure S7A).

In this study, statistically significant relationships between the opening size and harmonic, ultraharmonic, and broadband cavitation energies were detected (Figure 5C), consistent with our preclinical studies with mice and NHPs [33,48,49]. A recent study with MRgFUS in humans also showed the correlation between the subharmonic acoustic emission and the contrast-enhanced T1-weighted MR signal [23]. All subjects with BBBO showed overall increases in CD 20–30 s after the microbubble bolus injection (Figures 5A and 5B), indicating the onset of microbubble cavitation activity in the sonicated region. In some cases, the CD showed a high fluctuation before the major increase (harmonic CD in subject 2 and ultraharmonic CD in subject 4) or did not increase even after the flush (harmonic CD in subject 4). Compared to our preclinical studies with the same FUS transducer [33,34] where flat CD was usually observed before the injection in NHPs fixed by a stereotaxic frame, the baseline CD in this study was relatively unstable potentially due to motion. In addition, fluctuations in CD can also result from tissues, small air-bubbles in the coupling gel, or the membrane on the water cone [50]. The higher fluctuations observed in the harmonic and ultraharmonic CD profiles can be attributed to the measurements based on peak amplitude, whereas the broadband CD was determined by the averaged amplitude within a bandwidth. Additionally, spectral leakage from narrowband (harmonic and ultraharmonic) signals into the broadband may have occurred. An improved method for quantifying narrowband and broadband signals could be employed in future studies [51].

Harmonic and ultraharmonic CDs are typically associated with stable cavitation, characterized by the repetitive oscillation of microbubbles. Meanwhile, broadband CDs are more closely linked to inertial cavitation, which occurs when microbubbles collapse, leading to more violent mechanical effects. Due to these effects, monitoring of broadband CDs has been utilized to prevent tissue damage in mice [52,53] and non-human primates [33]. Similarly, efforts have been made to minimize the microbubble nonlinear behavior (i.e., subharmonic and ultraharmonic emissions) to reduce the possibility of damage in rabbits [54,55]. In our clinical study, BBBO was achieved without MRI-detectable damage in subjects 2 and 4, despite a 10–20 dB increase in ultraharmonic and broadband CDs observed. This suggests that not only stable but also stable-inertial and inertial cavitation was likely involved in facilitating BBBO [9,56]. Further studies are warranted to establish safe thresholds for ultraharmonic and broadband CD levels that effectively induce BBBO without causing tissue damage.

The reduction in SUVR was less pronounced in our study than in previous studies using an implantable FUS device [27] or using an MRgFUS system [16–19,26]. This difference may be attributed to the fact that we conducted a single session of treatment, while the prior studies involved 2–7 treatment sessions with larger treatment volumes. Nevertheless, we found significant decreases in asymmetry at the 2nd follow-up (Figure 7E and 7F), indicating a lower Aβ accumulation rate or elimination of amyloid in the treated side compared to the contralateral side. These asymmetry changes after FUS are consistent with findings from prior studies [16–19,25–27]. In addition, this lowered asymmetry SUVR across the treated frontal lobe and the hemisphere demonstrates the potential of FUS to exert holistic therapeutic effects beyond the treated region. When measured within the BBBO volumes in GM and WM (Figure 7D), the asymmetry did not show an apparent group-wise reduction possibly due to the small and variable BBBO volumes across the subjects. However, they correlated with the FUS treatment characteristics (i.e., BBBO size and CCDs) (Figures 7G and 7H). A larger BBBO or a higher harmonic CCD was related to the reduced Aβ accumulation in the treated region relative to the contralateral region. The observed decrease in SUVR asymmetry may suggest the effectiveness of the targeted treatment, potentially indicating a slowing or alteration in disease progression. Nevertheless, the small number of subjects in this pilot study warrants further investigation with larger cohorts to confirm these preliminary findings.

Preclinical studies have reported improved cognitive function after FUS [14,57,58] and a clinical study using MRgFUS has shown a cognitive improvement measured by the caregiver-administered neuropsychiatric inventory (CGA-NPI) [17]. However, the majority of clinical studies so far have reported non-significant changes in cognitive improvement following FUS-induced BBBO, examined by MMSE and ADAS-cog, evidencing no worsening of cognitive decline due to FUS [16,18,19]. Our MMSE results are also consistent with these findings.

A previous MRgFUS study reported significant increases in CSF T-Tau and CSF and plasma neurofilament light chain levels 1 week after MRgFUS and associated the increases with the T2* hypointensity findings in two patients [19]. In our study, although no group-wise changes in biomarker levels were found, the BBBO volume was significantly correlated with the increased EV levels of GFAP, Tau and pTau-181, 3 days after NgFUS without any abnormalities in MRI. The absence of a significant correlation between changes in Aβ biomarker levels and BBBO volume could be due to the small sample size or a weaker association with BBBO compared to other biomarkers, given Meng et al.’s study, which also reported no significant changes in Aβ biomarker levels [19]. The elevated levels of EV biomarkers indicate the release of proteins to the bloodstream by FUS, consistent with sonobiopsy behavior noted in prior studies [59,60]. As the BBBO volume also correlated with the reduced SUVR increase in the treated brain region, further investigation is warranted to discover the potential of FUS for clearing Alzheimer’s disease-related proteins from the brain to the bloodstream. The overall increased correlation of BBBO volume with the proteins in serum-derived EVs compared to serum levels alone indicates that EVs may be a more sensitive diagnostic tool for biomarker detection as a result of FUS-mediated BBBO. Mitochondria or endosome contamination was not accounted for in this study and may have influenced the absolute biomarker levels. While this study captures biomarker changes at a single time-point, extending our observations over multiple time-points would provide a more comprehensive understanding of biomarker release attributed to FUS.

Despite the promising findings of the study reported herein, there are several limitations, including a limited number of subjects, a single treatment at a single target location, inconsistency in BBBO volume, and targeting errors. This was a single-arm phase I trial with only six subjects and no control group. Despite the significant trends and differences observed in cavitation, biomarkers, and PET analysis, the results of this study should be interpreted with caution due to the small number of subjects. Furthermore, without a control group, the MMSE results might be influenced by placebo effects and should be interpreted with caution. Additionally, since multiple treatments have proven beneficial [26], evaluating the safety of regular NgFUS treatments is required. Future multi-arm phase II/III trials will not only involve a larger number of subjects and incorporate sham groups but will also include multiple NgFUS treatments to assess the safety of repeated sessions. Lastly, since our study included only mild-to-moderate AD patients, our results may not necessarily be generalizable to patients with lesser severity (pre-symptomatic), greater severity (severe disease), or those with various medical comorbidities.

To achieve more consistent BBBO volume across subjects, precise transducer positioning and accurate patient-specific simulations will be necessary in future studies. Based on our acoustic simulations, maintaining a transducer positioning error less than 3–5 mm in distance and 4–6° in angle is required to ensure an error margin of less than 10% in in-situ pressure. Robotics with neuronavigation guidance could be utilized to minimize manual positioning errors, while patient-specific fiducial markers could help reduce potential errors in registering the subject’s head to a virtual space. Moreover, we anticipate that the updated simulation pipeline will yield more accurate skull-induced attenuation estimates, given the robust correlation observed between the re-estimated attenuation and the resulting BBBO volume (Figure S8). Additionally, to further reduce inter-patient variability, we plan to implement real-time closed-loop feedback controllers based on cavitation metrics [61].

Furthermore, we plan to advance from our 2-D PAM to 3-D PAM using a matrix array probe, providing more comprehensive volumetric cavitation information. Additionally, we will further improve the mapping by employing skull-induced aberration correction.

Another limitation of our study is the small treated volume, considering that Alzheimer’s disease impacts broad regions of the brain [17,19]. To achieve more effective outcomes, our portable NgFUS system could adopt a larger volume treatment approach by utilizing a robotic arm, similar to a pre-clinical study by Leinenga et al [13]. In addition, a subject-specific hologram lens could be employed for a larger and constant focal size across subjects [62].

## Conclusions

The study presented herein demonstrates the safety and feasibility of transient and non-invasive BBBO in patients with Alzheimer’s disease using a portable NgFUS system. The BBBO volume showed linear correlations with the treatment dose (i.e., CCDs), the elevated level of biomarkers in serum-derived EVs, and the asymmetry SUVR changes. This low-cost and reliable technology may facilitate wider adoption of FUS treatment at the point of care for not only Alzheimer’s disease but also for several other neurological disorders.

## Materials and Methods

### Study Design

This study was a phase 1 clinical trial (NCT04118764) of six subjects for evaluating the safety and feasibility of NgFUS-mediated BBBO in patients diagnosed with mild-to-moderate Alzheimer’s disease. A power analysis prior to the trial showed that at least six subjects are needed to report a result with a significance level of 0.05, a power of 0.95, and an effective size of 4 (e.g., to detect a difference before and after FUS with a volume of 200 mm^3^ with a standard deviation of 50 mm^3^). The study was approved by the FDA and the Institutional Review Board (IRB) at Columbia University. After providing informed consent, participant eligibility was determined by the neurologist on the study based on the MRI and 18F-florbetapir PET scans, the participant and family interview, and clinical scales including MMSE, geriatric depression scale (GDS), and modified Hachinski ischemia scale (MHIS) (Table S1). Out of the ten subjects screened, four subjects were excluded due to low MMSE scores or the need for other medical treatment, and six subjects were enrolled in the study. The timeline of the study is presented in Figure 2. All subjects had baseline MRI and PET-CT scans 1–4 months before the treatment. For treatment planning, acoustic simulations were performed to estimate the skull-induced ultrasound attenuation and determine the FUS transducer output for each patient. On the day of treatment, the patient underwent one session of FUS sonication and post-treatment MRI was obtained approximately 2 h after the sonication to assess BBBO and safety. We aimed to establish a safety baseline with a single treatment session before introducing more complex protocols involving multiple sessions. Follow-up MRI scans were acquired 3 days after sonication to confirm BBB reinstatement and safety. Two follow-up PET scans were performed 3 weeks and 3 months after FUS for all subjects except Subject 1, who underwent follow-up PET scans at 3 days and 5 months, respectively. A 3-month follow-up period was deemed sufficient to evaluate acute and mid-term side effects, assuming that long-term adverse effects due to FUS were unlikely to occur more than 3 months post treatment. A follow-up MMSE was administered on the day of the 2nd follow-up PET. The timeline for each subject is listed in Table S6.

### NgFUS System

We used a single-element 250-kHz FUS transducer (H-231, Sonic Concepts) with a central opening, with guidance achieved using a neuronavigation system (Brainsight; Rogue Research) which was first tested in NHPs [32–34]. The FUS device was cleared by the FDA through an investigational device exemption (IDE G180140) for a first-in-human study at Columbia University. The −6 dB focal volume of the FUS beam was 6×6×49 mm^3^ with an axially-elongated ellipsoidal shape. For cavitation monitoring, either a single-element transducer for subjects 1–4 (Figure S11A) or a multi-element imaging array transducer for subject 5 and 6 (Figure S11B) was coaxially inserted in the central opening of the FUS transducer. A research ultrasound system (Vantage 256, Verasonics) was used for cavitation map acquisition. The transducer specifications and experimental parameters are listed in Table S7.

### Treatment Planning

The target location was selected at an amyloid positive region in the right frontal lobe based on the PET image. The initial FUS trajectory was determined by considering the focal size and the beam incidence angle relative to the skull (Figure 3) for more efficient acoustic energy delivery [38]. Patient-specific numerical simulations were employed for estimating the skull insertion loss of the acoustic pressure using the k-wave toolbox [63,64] and MATLAB (Figure 3B). Heterogeneous maps of the skull density and sound speed were obtained from the CT image acquired during screening (resolution: 0.6×0.6×1 mm^3^, Biograph64 mCT, Siemens), where the maximum sound speed and density were assumed to be *c* = 4000 m/s [65] and *p* = 1850 kg/m^3^ [66]. Skull absorption was also modelled based on the CT image with a maximum absorption value of 0.68 dB/cm at the working frequency, assuming a linear frequency dependency [66,67]. A 3-D acoustic pressure map was obtained from the linear acoustic simulation with a grid size of 1×1×1 mm^3^ (i.e., 6 points per wavelength) and a time step of 52.5 μs. The insertion loss *α* was determined by *α* = 1−*P*_skull_/*P*_freefield_, where *P*_skull_ is the maximum pressure within the brain obtained from a simulated acoustic map with skull insertion and *P*_freefield_ is the maximum pressure from a simulated map without the skull. More than 35 simulations were performed per subject considering the transducer positioning deviations (i.e., ±10 mm in distance and ±10° in angular deviation). The trajectory was also adjusted to avoid a large deviation of the insertion loss based on the simulation, and was used for FUS treatment as the planned trajectory. The estimated insertion loss along the planned trajectory (Table 2) was used for adjusting the sonication power to deliver the derated *in situ* pressure of 200 kPa.

### FUS Sonication

The dimensions of the portable FUS system required patients to have partial hair shaving at the right frontal scalp for optimal acoustic coupling between the subject’s head and the transducer (Figure S1A). The subject’s head was supported with the head and chin rest in a sitting position (Figure 3C), and the anatomical registration to the neuronavigation system was performed based on the facial landmarks (i.e., eyes, ears, and nose). The chin and head rest was used for subjects 2, 4, 5, and 6. The FUS transducer was positioned with the neuronavigation guidance to place the acoustic focus at the planned target in the right frontal lobe (Figure 3D). We determined the sonication parameters based on our previous simulations and pre-clinical studies [32,33]. The sonication parameters were as follows: derated peak-negative pressure, 200 kPa; mechanical index (MI), 0.4; center frequency, 0.25 MHz; pulse length, 10 ms; pulse repetition frequency, 2 Hz. treatment duration, 2 min. Microbubbles (0.1 mL/kg, Definity, Lantheus) were intravenously injected as a bolus starting at 3 s and finishing at 10–20 s after the start of the sonication, and followed with a saline flush. Approximately 50–75% of the microbubble bolus was introduced into circulation at the time of the flush due to the dead space within the catheter tubing. During the sonication, the frequency spectrum and cavitation dose were monitored (*N*=4), and the cavitation map with ultrasound B-mode image was also employed for the last two subjects (*N*=2) (Figure 3E).

### MRI and BBBO Quantification

Baseline (screening), post-FUS (day 0, 2 hr after FUS), and follow-up (day 3) MRI scans were acquired (Figure 2) using a 3-T MRI system (Signa Premier, GE). Safety MR scans were obtained during all three MRI sessions without any MR-contrast agent and included T2-weighted, T2-FLAIR, and SWI with parameters shown in Table S8. T1-weighted images with the gadolinium contrast agent (0.2 mL/kg, Dotarem®) were acquired for the confirmation of BBB opening and closing on day 0 and day 3, respectively. The post-contrast T1-weighted MRI was obtained 15–20 min after the gadolinium injection for increased sensitivity to detect BBBO [68,69]. BBBO on day 0 and closing on day 3 were confirmed by a neuro-radiologist.

The contrast-enhanced volume was quantified by subtracting the day-3 post-contrast T1-weighted MRI from the day-0 post-contrast T1-weighted MRI and thresholding the subtracted image. The threshold was automatically selected so that the mean intensity within the opening volume is significantly greater than that of the surrounding region with a confidence level of 98% assuming the intensity of the subtracted image follows a Gaussian distribution [34]. Evaluation of BBBO by tissue types was performed using the brain segmentation methods described in the Supplementary Methods.

### Blood Collection and Biomarker Measurement

Blood was collected from patients both prior to BBBO and 3 days post-BBBO to assess blood-based Alzheimer’s disease biomarker detection as a result of FUS from both serum and serum-derived EVs. All subjects had blood drawn immediately 1–2 hours prior to the treatment (i.e., baseline) and 3 days after treatment. Serum was isolated after centrifugation of whole blood at 9.4 rcf for 5 min at 4 ℃, and serum-derived EVs were isolated using an exosome precipitation solution according to the manufacturer’s published protocol (ExoQuick, Systems Biosciences, Palo Alto, CA). A Luminex multiplex assay was used to quantify proteins in serum and in isolated serum-derived EVs (Luminex Corp., Austin, TX). Single pro-cartaplex kits (ThermoFisher Scientific) were purchased and combined to make a custom multiplex panel for analysis. The biomarker levels of subject 1 were not acquired properly because of mishandling of the blood specimen.

### PET/CT

PET/CT scans (CT: no contrast, axial plane, 4 mm section thickness, 4mm section interval) were acquired with a clinical PET scanner (Biograph64-mCT; Siemens) and with ^18^F-Florbetapir tracer at 10mCi (Amyvid®; PETNET Solutions). PET/CT scans were acquired 32 to 107 days prior to treatment, 3 to 29 days after treatment as the 1st follow-up time-point, and 82 to 164 days after treatment as the 2nd follow-up time-point (Table S6). The Aβ load was quantified from PET scans as SUVR, using the cerebellar GM as the reference region [70]. For region-specific amyloid analysis, MRI and PET images were registered to the MNI space and automatically segmented by tissue types. To investigate changes in Aβ from a localized region at the site of BBBO to extended regions, three areas were analyzed: BBBO volumes in the GM and WM (SUVR_BBBO*_), the treated right frontal lobe (SUVR_FL_), and the treated right hemisphere (SUVR_H_). The asymmetry SUVR (Asym.SUVR) was measured by dividing the average SUVR in the treated region by that in the contralateral region to monitor the relative progression of Aβ. Changes in SUVR and asymmetry at the 1st and 2nd follow-ups compared to baseline were quantified (Figure 7). SUVR in Centiloid scale was calculated using established methods [71–73], in order to allow standardized comparison with other studies. Details of PET analysis are presented in the Supplementary Methods.

### Cavitation Dose and Cavitation Mapping

For cavitation monitoring, PCD was used for subjects 1–4 and PAM was employed for subjects 5 and 6. Device specification and parameters are listed in Table S7. The CD was obtained from the 3rd to 6th harmonic/ultrahamonic frequencies. We computed the CD with harmonic (CD_h_), ultraharmonic (CD_u_), and broadband frequencies (CD_b_) as described in the Supplementary Methods. The CCD was obtained by summing the normalized CD acquired after the microbubble flush and converting it to the logarithmic scale.

The cavitation map for each burst was reconstructed in real time from the 64-channel RF data by using the coherence-factor-based PAM implemented on a GPU (RTX A6000, NVIDIA). The final cavitation maps were obtained by averaging the acoustic energy maps for the bursts after the microbubble injection and masking them with the segmented brain volumes obtained from the MR images. More information on PAM implementation can be found in our previous study [34].

The BBBO volumes quantified in MR images were registered with the cavitation maps based on the tracked coordinate of the focus by the neuronavigation system and also based on ultrasound B-mode image that delineated the skin and skull; the registered B-mode (or cavitation map) and MRI slice are presented in Figure S12. We evaluated the predictive capability of each pixel in the cavitation image for detecting BBBO using the pixel-wise correlation. The AUC of ROC and PR curves were calculated following the methods described in our previous study [34].

### Targeting Accuracy and Precision

The planned target/trajectory of FUS was determined in the planning step before treatment and the treated target/trajectory was sampled during the FUS sonication on the neuronavigation system. Transducer positioning errors were measured by the distance and angle differences between the planned and treated target trajectories to assess the accuracy in the manual placement of the transducer. To evaluate the targeting accuracy of BBBO, the Euclidean distance between the BBBO centroid and the simulated focus was measured for each subject. The subject movement was obtained from the tracked location of the FUS focus which was recorded over time during the sonication by the neuronavigation system.

### Post Hoc Simulation

During the retrospective analysis of data, we found that there was a registration error between CT and MR volumes (1–7 mm). Additionally, there were differences between the treated trajectory for sonication and the planned trajectory for the acoustic simulation before treatment, due to the transducer positioning error. We re-simulated the acoustic pressure fields with the corrected registration and the trajectory. The pressure field (Figure S7), attenuation, and derated peak pressure (Table S5) were obtained with the corrected trajectory and registration. The derated peak pressure *P̂* was calculated by *P̂* = *P*/(1 − *⍺*) · (1 − *⍺̂*) where *P* is 200 kPa and *⍺* and *⍺̂* are the original and the newly obtained insertion loss values, respectively.

### Statistical Analysis

Statistical analysis was performed in MATLAB (Mathworks). Linear regression analysis was used to evaluate the correlations of the CCDs (*N*=4), biomarker levels (*N*=5), asymmetry SUVR increase, and the simulated maximum pressure (*N*=5) with the contrast-enhanced volume, as well as the correlation between the SUVR asymmetry and CCDs (*N*=4). R-squared and p values were obtained from the regression for the statistical analysis. Pixel-wise correlation between the cavitation map and the BBBO was measured by the AUC of ROC curve and the AUC of PR curve after combining data sets from subject 5 and 6, as described in the previous study [34]. MMSE scores of the subjects were compared with those of ADNI subjects by using unpaired t-test. Changes in SUVR and the asymmetry between different time-points were analyzed using paired t-test.

## Data Availability

All study data are included in the main text and/or supplementary materials.

## Abbreviations

Aβ: Amyloid beta
AD: Alzheimer’s disease
ADNI: Alzheimer’s disease neuroimaging initiative
AE: Adverse event
BBB: Blood-brain barrier
BBBO: Blood-brain barrier opening
CCD: Cumulative cavitation dose
CGA-NPI: Caregiver-administered neuropsychiatric inventory
CT: Computed tomography
EV: Extracellular vesicle
FDA: U.S. Food and Drug Administration
FLAIR: Fluid-attenuated inversion recovery
FUS: Focused ultrasound
GM: Gray matter
GPU: Graphics processing unit
IRB: Institutional review board
MMSE: Mini-mental state examination
MRI: Magnetic resonance imaging
MR: Magnetic resonance
MRgFUS: Magnetic resonance-guided focused ultrasound
NHP: Non-human primate
NgFUS: Neuronavigation-guided FUS
PAM: Passive acoustic mapping
PCD: Passive cavitation detection
PET: Positron emission tomography
PR: Precision-recall
ROC: Receiver operating characteristic
SAE: Adverse event
SUVR: Standard uptake value ratio
SUVR_BBBO*_: Standard uptake value ratio measured within BBBO volume in the gray and white matter
SUVR_FL_: Standard uptake value ratio measured within the treated frontal lobe
SUVR_H_: Standard uptake value ratio measured within the treated hemisphere
SWI: Susceptibility-weighted imaging

## Acknowledgments

The authors wish to thank Maria F. Murillo, Alexander Berg, Rebecca L. Noel, and Nancy Kwon for support and insightful discussion.

## Contributions

ANP, EEK, and LSH initially developed the concept of the study;

SB, KL, RJ, SJG, OY, FNT, ANP, DK, LSH, and EEK conducted the FUS treatments;

LSH performed the neurological examinations;

SB processed and analyzed the cavitation, patient motion, and MRI data;

OY, KL, SJG, and FNT performed the numerical simulations;

KL and RJ processed and analyzed PET data;

AKS, AJB, SB, and FNT processed and analyzed blood biomarker data;

SB, RJ, SJG, KL, AJB, and EEK discussed and reviewed all the data;

KL, DK, ANP, and RJ assisted in patient recruitment;

ANP, KL, RJ, and SB prepared technical and regulatory documentation of the device;

EEK acquired funding and provided resources for the study as a principal investigator;

EEK and LSH supervised the clinical study;

SB wrote the original manuscript and visualized the data;

SB, KL, ANP, RJ, LSH, EEK, AJB, and SJG and edited the manuscript;

All authors reviewed and approved the final manuscript.

## Competing interests

Some of the work presented herein is supported by patents optioned to Delsona Therapeutics, Inc. where EEK serves as co-founder and scientific adviser. SB, ANP, RJ, KL, SJG, OY, FNT, DK, AKS, AB, and LSH declare no conflict of interest.

## Notes

### Clinical Trial

NCT04118764, IDE-G180140

### Clinical Protocols

https://clinicaltrials.gov/study/NCT04118764

### Funding Statement

National Institutes of Health grant R01AG038961 (EEK)
National Institutes of Health grant R01EB009041 (EEK)
Focused Ultrasound Foundation (EEK)

### Author Declarations

Institutional Review Boards of Columbia University gave ethical approval for this work

### Summary of Updates

We had a major revision. Specifically, we have included new paragraphs in the Discussion section regarding cavitation activity, pressure estimation errors, and future plans to address the limitations of the study. Additionally, we have added to the Supplementary Methods and Discussion to provide a more detailed explanation of our methods and analysis.

