## Supplementary for "Transcranial Blood–Brain Barrier Opening in Alzheimer’s Disease Patients Using a Portable Focused Ultrasound System with Real-Time 2-D Cavitation Mapping"

**This PDF file includes:**

Supplementary Figures S1 to S12
Supplementary Tables S1 to S8
Supplementary Methods
Supplementary Discussion

**Other Supplementary Materials for this manuscript include the following:**

Movie S1

### Supplementary Figures

**A** **B**

Please contact the corresponding author to request access to this figure.

**Figure S1. Partial shaving and transducer contact area.**

**(A)** The hair-shaved area (white dashed line) and transducer contact area (black dashed line) with a diameter of 50 mm for the FUS sonication at the right frontal lobe. **(B)** The shaved area (white dashed line) of subject 6 was fully covered by parting hair after the treatment.

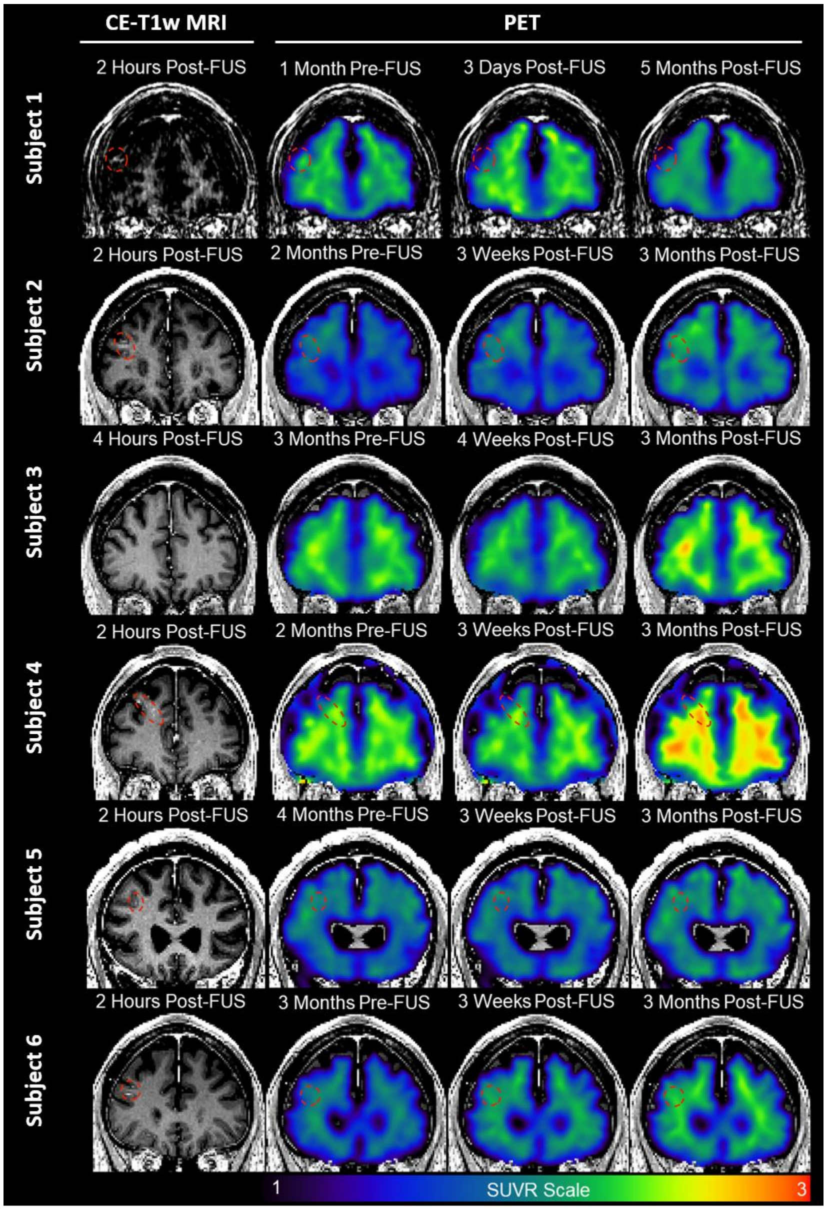


**Figure S2. Contrast-enhanced T1-weighted MRI and PET images of six subjects.**
All scans are registered to the Montreal Neuroimaging Institute (MNI) space. The first column shows representative planes of respective subjects' MRI scans after treatment with contrast enhancement indicating BBBO marked by a red dashed circle. Subject 3 had no BBBO. The second to fourth columns included the same plane of MRI overlaid with PET SUVR (scale from 1 to 3) at pre-FUS, 1st follow-up post-FUS, and 2nd follow-up post-FUS, respectively. Methods for PET imaging processing are presented in the Supplementary Methods. The time-point of each scan is denoted above each image. The voxel intensity of PET images in SUVR scale was calculated by dividing the tracer uptake at each voxel by the average tracer uptake within the reference region of cerebellar gray matter.

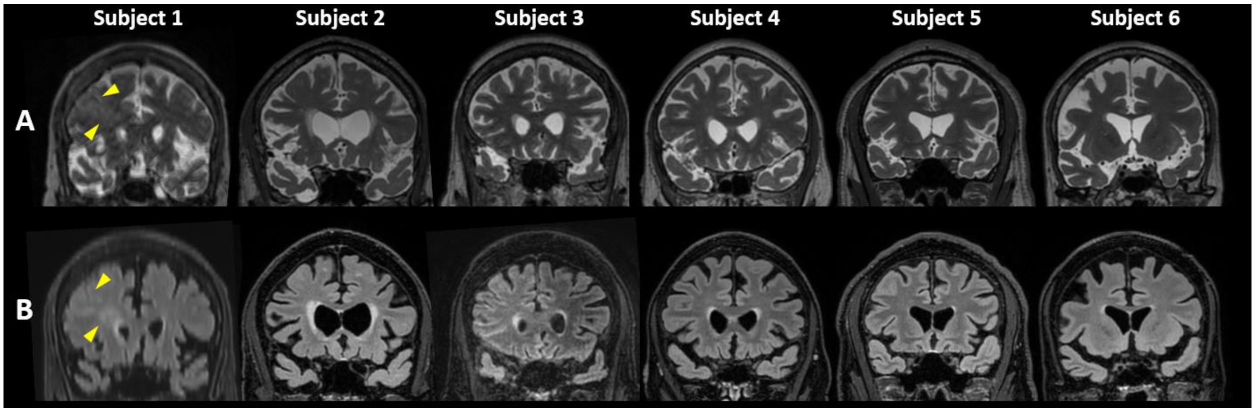


**Figure S3. Safety MRI obtained on day 3 post-FUS**

**(A)** T2-weighted and **(B)** T2 fluid-attenuated inversion recovery (FLAIR) MRI on day 3 for safety assessment after FUS. Subject 1 exhibited signs of edema at the targeted region in both T2-weighted and T2-FLAIR images (arrowheads), which was resolved in the follow-up scans. Other subjects had no detectable brain imaging abnormalities found in the safety scans.

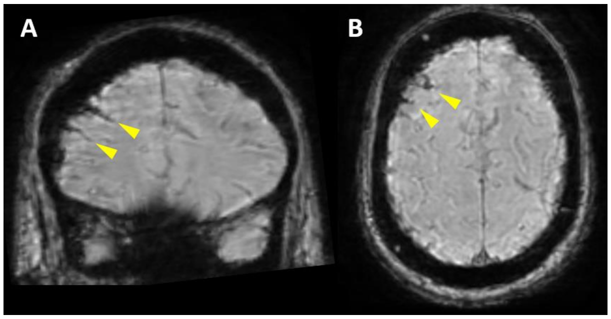


**Figure S4. Susceptibility-weighted imaging (SWI) in subject 1 in the (A) coronal and (B) axial planes, indicating the superficial subarachnoid hemorrhage.**

This asymptomatic finding was resolved in 15 days.

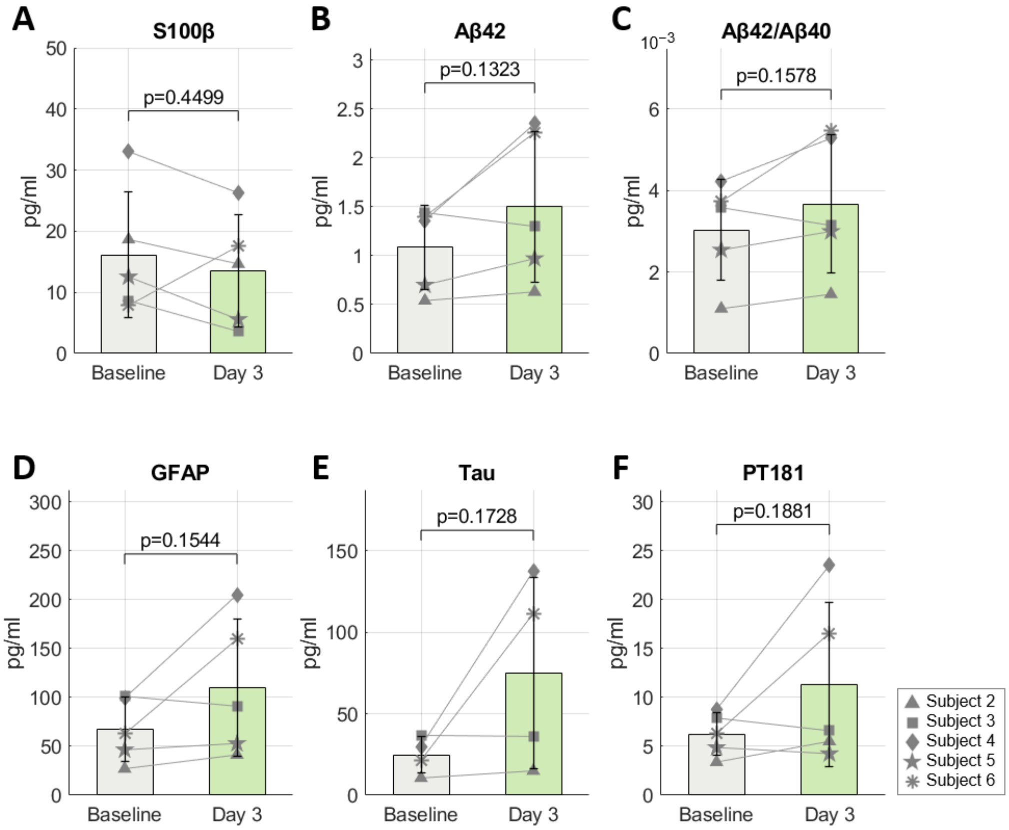


**Figure S5. Serum and extracellular vesicle biomarker analysis.**

Biomarker levels of (**A**) S100β in serum, (**B**) Aβ42, (**C**) Aβ42/Aβ40, (**D**) GFAP, (**E**) Tau, and (**F**) pT181 in extracellular vesicles (EV) were measured at the time points of baseline (1–2 hours before NgFUS) and day 3 (72 hours after NgFUS). No significant group-wise changes (paired t-test) were observed, possibly because of the large variation in BBBO volume across subjects.

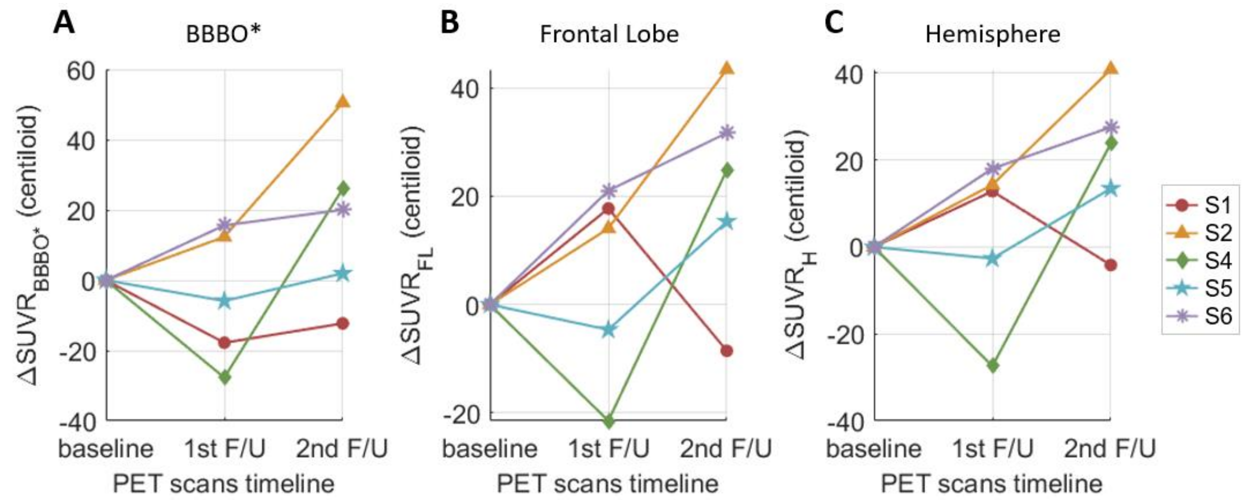


**Figure S6. Changes in the standard uptake value ratio (SUVR) in Centiloid units.** SUVR measured within the blood-brain barrier opening (BBBO) volume in the gray and white matter ($\Delta SUVR_{BBBO*}$), the right frontal lobe ($\Delta SUVR_{FL}$), and the right hemisphere ($\Delta SUVR_{H}$), at the 1st and the 2nd follow-ups compared to the baseline. Subject 3 was excluded due to the absence of BBBO. (BBBO*: BBBO in the gray and white matter)

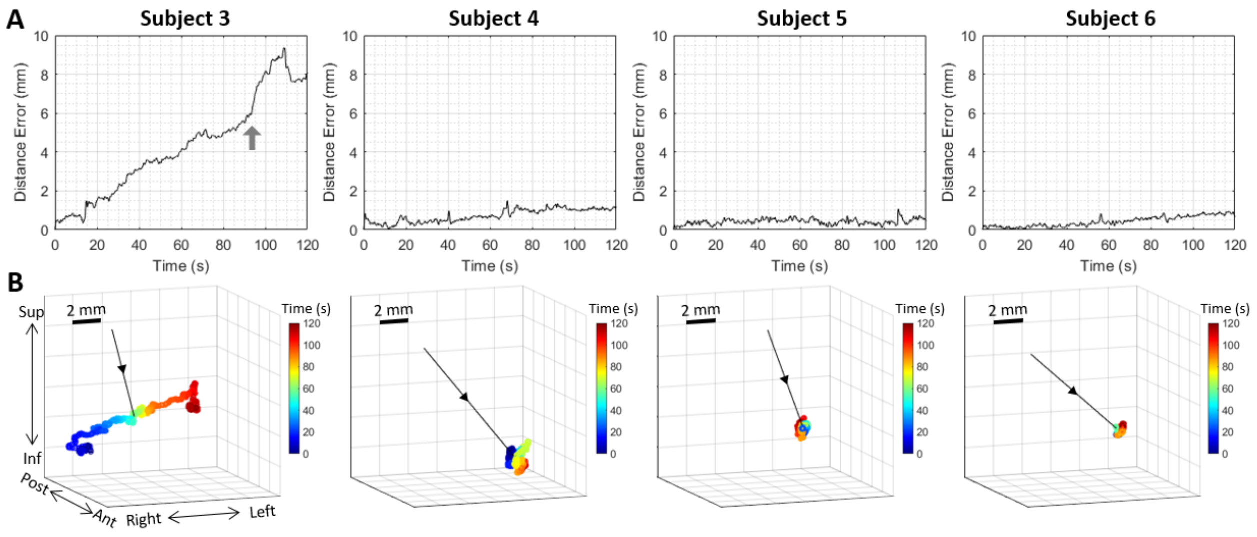


**Figure S7. Subject movement tracking during the treatment.**

**(A)** Distance from the initial position over time presents the FUS focal shift due to the unrestrained subject movement. The sudden large shift at $t$ = 90 s in subject 3 (marked with a gray arrow) coincided with the cavitation dose reduction of subject 3 in Figure 5A. **(B)** Tracked 3-D position of the FUS focus relative to the subject's head during the 2-min treatment. The black lines in (B) represent the FUS beam axis, and the time is color-coded. In subject 3, the focus was shifted to the left mainly due to the subject motion, while other subjects showed a limited movement of less than 2 mm during the treatment.

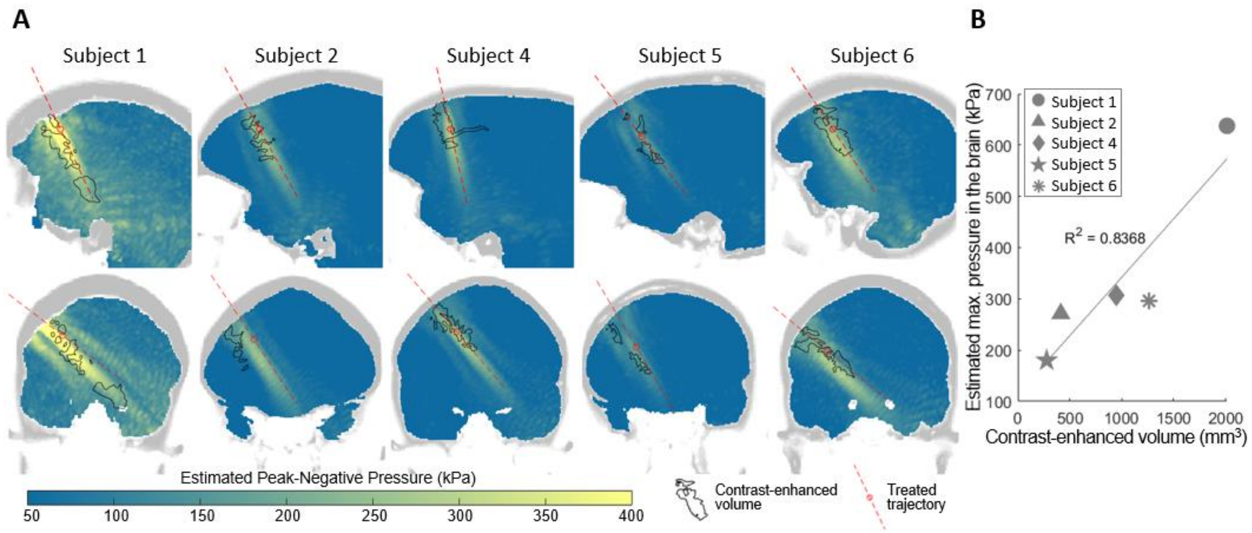


**Figure S8. Post-hoc simulation Results**

**(A)** Max-projection of the pressure map obtained from the post hoc simulation in the sagittal (top) and the coronal views (bottom) and **(B)** the correlation between the maximum pressure and contrast-enhanced volume. The map in (A) shows higher pressure for subjects with larger opening volume (black contour). In subject 1, the linear hyperintense region (yellow) in the coronal map shows a refraction of the beam toward the inferior compared to the incident trajectory (red dashed line) and overlaps with the contrast-enhanced volume (black contour). The estimated maximum pressure linearly correlates with the opening volume across the subjects with an R-squared value of 0.84 in (B).

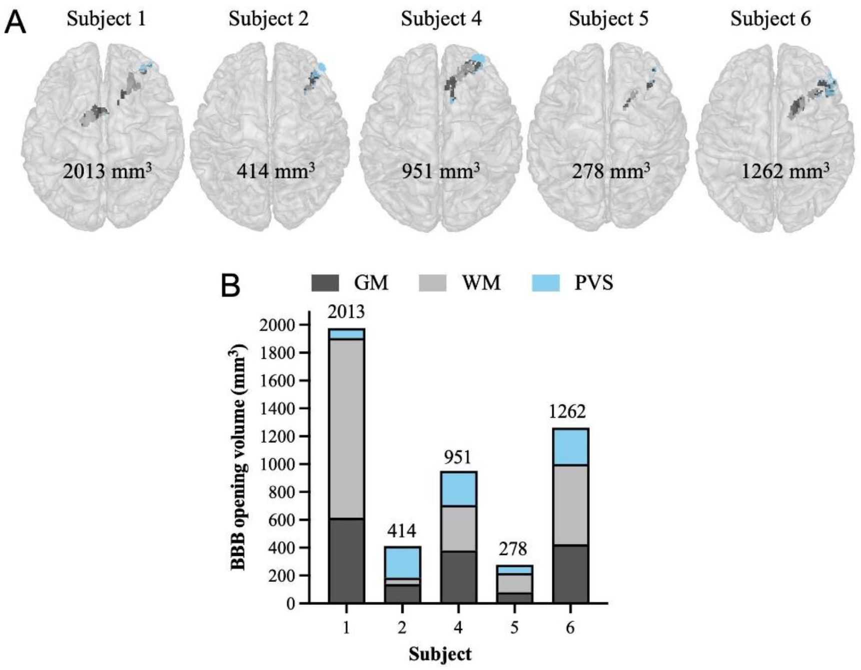


**Figure S9. BBBO volumes with segmentation by tissue types.**

BBBO volumes **(A)** overlaid on MNI-registered brains and **(B)** plotted in proportions of gray matter (GM), white matter (WM), and perivascular space (PVS). On average, 33.23% ± 4.39% of the BBBO volumes was in the GM, 41.00% ± 9.90% was in the WM, and 25.26% ± 18.56% was through vessels lining the PVS**.**

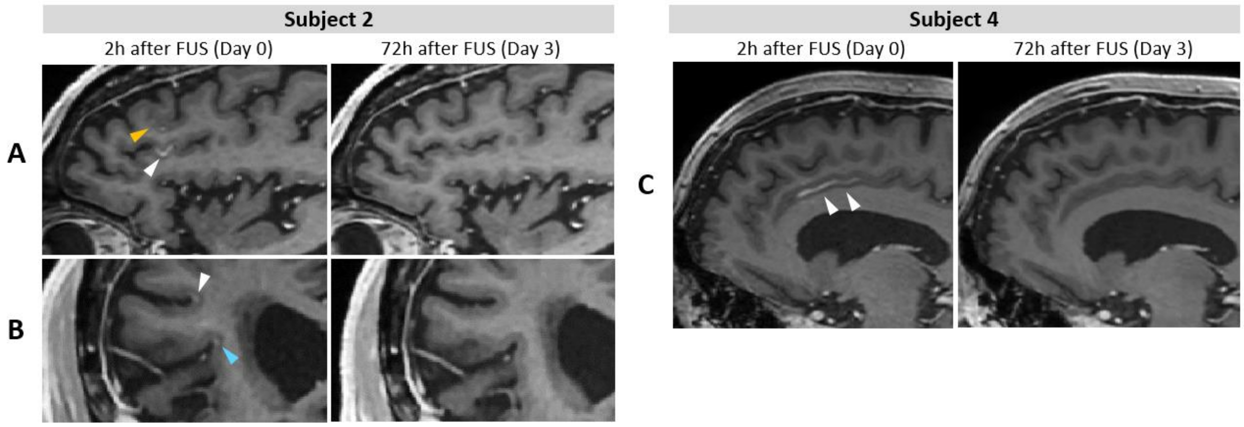


**Figure S10. Contrast enhancement along the sulci in subjects 2 and 4 in T1-weighted MRI.** **(A, B)** Contrast enhancement was observed 2h after FUS along the middle frontal sulcus (yellow arrowhead), the inferior frontal sulcus (white arrowhead), and the Sylvian sulcus (blue arrowhead) in subject 2 in (A) the sagittal and (B) coronal views. **(C)** Contrast enhancement was observed 2h after FUS along the cingulate sulcus (white arrowheads) in subject 4 in the sagittal view (left panel) and was not visible 72 h after FUS (right panel), indicating the reinstatement of the BBB or blood-CSF barrier.

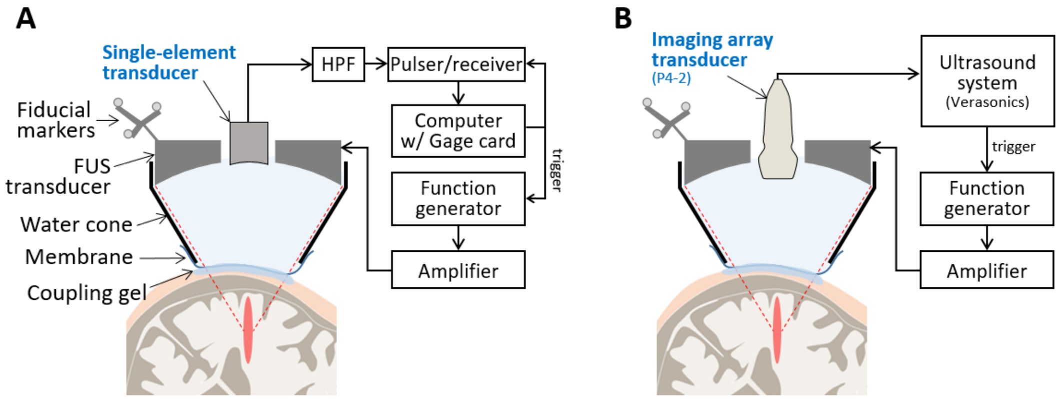


**Figure S11. FUS transducer with cavitation monitoring setup with (A) a single-element PCD and (B) an imaging array transducer for PAM.**

The FUS transducer was placed onto the subject's head and coupled with water and acoustic gel. The single-element transducer was used to measure cavitation doses in subjects 1–4, while the imaging array was employed to obtain both cavitation doses and cavitation maps in subjects 5 and 6. Specifications of these devices can be found in Table S5.

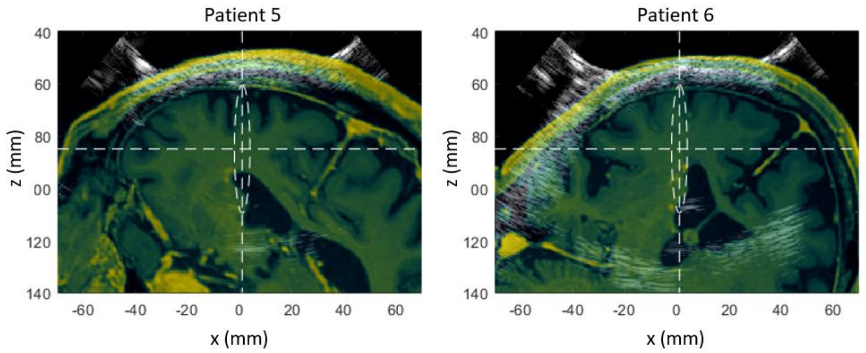


**Figure S12. Registration results between the ultrasound B-mode image (gray) and MRI slice (color) in subjects 5 and 6.**

The MRI slice aligned with the ultrasound imaging plane was obtained from the neuronavigation system for the spatial comparison between the cavitation map and the BBBO volume in MRI. Additional manual adjustments were made for more accurate registration between the two images based on the skull lines visible both in the B-mode and MRI images.

### Supplementary Tables

**Table S1. Inclusion and exclusion criteria for enrollment**

| Inclusion criteria | Exclusion criteria |
|---|---|
| • Age > 50<br>• Ability to provide informed consent<br>• Diagnosis of AD or MCI due to AD<br>• MMSE score between 12 and 26<br>• MHIS score ≤ 4<br>• GDS score ≤ 6<br>• PET scan confirming amyloid plaque load | • Prior brain surgery, including deep brain stimulation.<br>• Moderate or severe uncontrolled hypertension.<br>• Contraindication or hypersensitivity to MRI, MRI contrast agents, and microbubbles.<br>• Inability to comply with the procedures of the protocol, including follow-up MRI scans.<br>• Metallic implants.<br>• Abnormal coagulation profile or history of stroke or cardiovascular disease.<br>• Severe brain atrophy, tumor, space-occupying lesions, meningeal enhancements, or intracranial hypotension.<br>• History of seizure disorder. History of asthma or allergies to food or medication with significant symptoms in past 3 years.<br>• Pregnancy or lactation.<br>• Impaired renal function with estimated glomerular filtration rate (eGFR) < 30 mL/min/1.73m$^2$<br>• Active gingivitis, herpes simplex, hepatitis, tuberculosis, and minor skin or respiratory infections.<br>• Active infection/inflammation or acute/chronic hemorrhages. |

AD: Alzheimer's Disease, MCI: Mild Cognitive Impairment, MMSE: Mini-Mental State Examination, MHIS: Modified Hachinski Ischemia Scale, GDS: Short form Geriatric Depression Scale

**Table S2. Individual Subject's SUVR and asymmetry SUVR values**

| Subject# | Region | SUVR | | | Asym.SUVR | | |
|---|---|---|---|---|---|---|---|
| | | Baseline | 1st F/U | 2nd F/U | Baseline | 1st F/U | 2nd F/U |
| | BBBO* | 1.666 | 1.569 | 1.599 | 0.986 | 0.89 | 0.978 |
| 1 | FL | 1.871 | 1.968 | 1.825 | 0.987 | 0.984 | 0.979 |
| | H | 1.815 | 1.885 | 1.793 | 0.986 | 0.981 | 0.98 |
| | BBBO* | 1.813 | 1.88 | 2.087 | 1.1 | 1.094 | 1.085 |
| 2 | FL | 1.618 | 1.694 | 1.854 | 1.037 | 1.03 | 1.019 |
| | H | 1.558 | 1.636 | 1.781 | 1.01 | 1.012 | 1 |
| | BBBO* | 2.202 | 2.052 | 2.344 | 1.006 | 0.976 | 0.997 |
| 4 | FL | 1.95 | 1.833 | 2.085 | 1.04 | 1.012 | 1.013 |
| | H | 1.976 | 1.827 | 2.106 | 1.032 | 1.016 | 1.02 |
| | BBBO* | 1.383 | 1.351 | 1.394 | 0.849 | 0.879 | 0.859 |
| 5 | FL | 1.797 | 1.771 | 1.88 | 1.006 | 0.992 | 0.998 |
| | H | 1.723 | 1.709 | 1.797 | 1.002 | 0.992 | 0.996 |
| | BBBO* | 1.57 | 1.654 | 1.678 | 0.981 | 0.995 | 0.967 |
| 6 | FL | 1.551 | 1.665 | 1.723 | 1.036 | 1.038 | 1.022 |
| | H | 1.473 | 1.571 | 1.623 | 1.012 | 1.009 | 1.001 |

SUVR: standard uptake value ratio, Asym.: asymmetry, BBBO*: BBBO volume in gray and white matter, FL: frontal lobe, H: hemisphere, F/U: follow-up. The 1st and 2nd F/Us were 20±9 days and 103±30 days after focused ultrasound treatment, respectively. Subject 3 was excluded due to the absence of BBBO.

Participants' MMSE scores compared to ADNI comparison patients

**Table S3. MMSE Scores**

| | All participants | Participants with BBB opening | ADNI comparison patients[**] |
|---|---|---|---|
| N (Sex) | 6 (2M, 4F) | 5 (2M, 3F) | 33 (14M, 19F) |
| Age | 70±7 | 70 ± 8 | 74±7 |
| Baseline MMSE | 18.8±3.2 | 19.6 ± 3.0 | 22.0±2.8 |
| Follow-up MMSE[*] | 16.3±4.4 | 17.8 ± 3.3 | 19.2±3.2 |
| Changes | −2.5±2.9 (p=.79) | −1.8±2.7 (p=.45) | −2.8±2.8 |

MMSE: Mini-Mental State Examination, ADNI: Alzheimer's disease neuroimaging initiative.

Age, MMSE scores, and changes are expressed as mean ± standard deviation

[*]Follow-up MMSE was 103±30 days after treatment and 189±37 days after baseline MMSE.

[**]Patients (N=33, 19F, 14M) from the ADNI database (https://adni.loni.usc.edu/) with MMSE administered 6 months apart were selected by matching to each study participant by baseline MMSE score, age, and/or gender

**Table S4. Individual Subject's MMSE Scores**

| Subject # | Baseline MMSE | Follow-Up MMSE* | Change in MMSE |
|---|---|---|---|
| 1 | 18 | 18 | 0 |
| 2 | 20 | 22 | +2 |
| 3 | 15 | 9 | -6 |
| 4 | 21 | 19 | -2 |
| 5 | 24 | 18 | -6 |
| 6 | 15 | 12 | -3 |

*Follow-up MMSE was administered 103±30 days after FUS and 189±37 days after baseline MMSE.

**Table S5. Re-estimated skull-induced attenuation of FUS beam and the derated pressure obtained through the post-hoc simulation**

| Subject # | 1 | 2 | 3 | 4 | 5 | 6 |
|---|---|---|---|---|---|---|
| Re-estimated skull-induced attenuation* | 0.49 | 0.62 | - | 0.57 | 0.73 | 0.63 |
| Estimated peak pressure** (kPa) | 638 | 271 | - | 307 | 180 | 296 |

* Obtained by the post hoc acoustic simulation based on the treated target trajectory.
** Peak acoustic pressure within the brain estimated by the post hoc simulation.

**Table S6. MMSE and $^{18}$F-Florbetapir-PET time points for each subject**

| Subject # | Baseline MMSE, baseline PET, and CT | 1st F/U PET | F/U MMSE and 2nd F/U PET |
|---|---|---|---|
| 1 | 32 days pre-FUS | 3 days post-FUS | 164 days post-FUS |
| 2 | 68 days pre-FUS | 23 days post-FUS | 91 days post-FUS |
| 3 | 55 days pre-FUS | 29 days post-FUS | 98 days post-FUS |
| 4 | 76 days pre-FUS | 24 days post-FUS | 93 days post-FUS |
| 5 | 119 days pre-FUS | 24 days post-FUS | 91 days post-FUS |
| 6 | 107 days pre-FUS | 21 days post-FUS | 82 days post-FUS |

**Table S7. Device specifications and parameters**

| | | |
|---|---|---|
| FUS | Transducer model | H-231, Sonic Concepts |
| | Center frequency of the transducer | 0.25 MHz |
| | Outer/inner diameter of the transducer | 110/44 mm |
| | Radius of curvature of the transducer | 110 mm |
| | Focal size of the transducer | 6 mm × 6 mm × 49 mm |
| | Amplifier | A150 or A075, E&I |
| | Water degassing system | WDS105+, Sonics Concepts |
| Passive cavitation detection (PCD) for cavitation dose monitoring (subjects 1–4) | Transducer model | 1.7MHz/1.25″, ndtXducer |
| | Number of transducer element | 1 |
| | Diameter of the transducer | 32 mm |
| | Focal depth of the transducer | 114 mm |
| | Acquisition system | High-pass filter (HPF) (ZFHP-0R60-S+, Mini-circuits) and pulser/receiver (DPR300, JSR Ultrasonics) |
| | Sampling frequency | 50 MHz |
| Passive acoustic mapping (PAM) for both cavitation dose and mapping (subject 5 and 6) | Transducer model | P4-2, ATL |
| | Number of transducer elements | 64 |
| | Transducer center frequency | 2.5 MHz |
| | Transducer aperture size | 19.2 mm |
| | Acquisition system | Vantage 256, Verasonics |
| | Sampling frequency | 10 MHz |

**Table S8. MRI sequence parameters.** For T1, T2, and T2-FLAIR MRI, axial sequences were used for subject 1 and coronal sequences were used for other subjects.

| Sequence Name | T1 AXIAL | T2 AXIAL | T2 FLAIR AXIAL | SWI AXIAL |
|---|---|---|---|---|
| Repetition time (ms) | 8.44 | 3002 | 9000 | 40.04 |
| Echo time (ms) | 3.59 | 80.73 | 89.6 | 35.95 |
| Number of averages | 1 | 2 | 1 | 0.70 |
| Flip angle (°) | 11 | 90 | 160 | 15 |
| In-plane resolution (mm) | 0.5×0.5 | 1×1 | 0.47×0.47 | 0.86×0.86 |
| Slice thickness (mm) | 0.8 | 1 | 3 | 0.89 |

| Sequence Name | T1 CORONAL | T2 CORONAL | T2 FLAIR CORONAL |
|---|---|---|---|
| Repetition time (ms) | 7.44 | 2500 | 7500 |
| Echo time (ms) | 3.11 | 80.92 | 93.93 |
| Number of averages | 0.7 | 1 | 1 |
| Flip angle (°) | 11 | 90 | 90 |
| In-plane resolution (mm) | 0.41×0.41 | 0.82×0.82 | 0.41×0.41 |
| Slice thickness (mm) | 0.8 | 1 | 0.99 |

### Supplementary Methods

**Brain Segmentation for Region-Specific Analysis of MRI and PET**

To evaluate the BBBO by tissue types and investigate region-specific amyloid changes, all regions of interest were automatically created to reduce operator dependence. MRI and PET images were registered to the Montreal Neuroimaging Institute (MNI) space and automatically segmented by tissue types—gray matter (GM), white matter (WM), and perivascular space (PVS)—as well as into specific brain regions such as the frontal lobe (FL) and hemispheres (H) [1-3]. Image processing software used included Clinica [4], SPM12 [5], Mango [6], and FSL [7]. Probability maps for GM, WM, and PVS were obtained using SPM's unified segmentation algorithm and were registered to the MNI space using a diffeomorphic registration algorithm [5]. Binary masks were then created using a tissue probability cutoff of 0.5. The BBBO volume in the GM and WM (i.e., BBBO*) was obtained by combining the quantified BBBO mask with the segmented GM and WM masks.

The tissue-type-specific BBBO volumes in Figure S9 were determined using the GM, WM, and PVS binary mask. The region-specific SUVR and Asym.SUVR values in Figure 7 (e.g., $SUVR_{BBBO*}$, $SUVR_{FL}$, and $SUVR_{H}$) were obtained using the BBBO*, frontal lobe, and hemisphere masks, respectively, within the GM and WM.


[1] Rolls ET, Joliot M, Tzourio-Mazoyer N. Implementation of a new parcellation of the orbitofrontal cortex in the automated anatomical labeling atlas. Neuroimage. 2015; 122: 1–5.

[2] Fonov V, Evans AC, Botteron K, Almli CR, McKinstry RC, Collins DL. Unbiased average age-appropriate atlases for pediatric studies. Neuroimage. 2011; 54: 313–27.

[3] Rolls ET, Huang CC, Lin CP, Feng J, Joliot M. Automated anatomical labelling atlas 3. Neuroimage. 2020; 206.

[4] Routier A, Burgos N, Díaz M, et al. Clinica: An Open-Source Software Platform for Reproducible Clinical Neuroscience Studies. Front Neuroinform. 2021; 15.

[5] Ashburner J, Friston KJ. Unified segmentation. Neuroimage. 2005; 26: 839–51.

[6] Lancaster JL, Laird AR, Eickhoff SB, Martinez MJ, Mickle Fox P, Fox PT. Automated regional behavioral analysis for human brain images. Front Neuroinform. 2012; 6.

[7] Jenkinson M, Beckmann CF, Behrens TEJ, Woolrich MW, Smith SM. FSL. Neuroimage [Internet]. 2012; 62: 782–90. Available at: https://linkinghub.elsevier.com/retrieve/pii/S1053811911010603

**PET Imaging Processing**

The Aβ load was quantified from PET scans as standardized uptake value ratio (SUVR), using the cerebellar gray matter as the reference region. Supplementary Figure S2 shows PET scans in SUVR scale, calculated as follow:

$$SUVR(x, y, z) = I(x, y, z)/\bar{I}_{\mathrm{ref}}$$

where $SUVR(x, y, z)$ represents the intensity of PET image at each voxel $(x, y, z)$ in SUVR scale, $I(x, y, z)$ is the tracer uptake at each voxel, and $\bar{I}_{\mathrm{ref}}$ is the average tracer uptake within the reference region of cerebella gray matter specific to each subject.

Asymmetry SUVR in Figure 7 was obtained by dividing the average SUVR in the region of interest (ROI) by the average SUVR in the contralateral ROI, as follows:

$$Asym. SUVR = \overline{SUVR}_{\mathrm{ROI}}/\overline{SUVR}_{\mathrm{contralateral}}$$

where $\overline{SUVR}_{\mathrm{ROI}}$ and $\overline{SUVR}_{\mathrm{contralateral}}$ are the average SUVR within the ROI (BBBO, right frontal lobe, or right hemisphere within GM and WM) and that within the contralateral ROI (mirrored BBBO, left frontal lobe, or left hemisphere within GM and WM), respectively.

**Cavitation Dose Analysis**

Cavitation dose (CD) was obtained from the frequency spectrums of the received cavitation signal during the 2-min FUS. We computed the CD with harmonic ($CD_h$), ultraharmonic ($CD_u$), and broadband frequencies ($CD_b$) as follows based on our previous studies [1,2]:

$$\mathrm{CD_h} = \sqrt{\sum_{n=3}^{6} |P_{\mathrm{h},n}|^2}\ ,$$

$$\mathrm{CD_u} = \sqrt{\sum_{n=3}^{6} |P_{\mathrm{u},n}|^2},$$

$$\text{and } \mathrm{CD_b} = \sqrt{\sum_{n=3}^{6} |\bar{A}_n|^2},$$

where $P_{\mathrm{h,n}}$ and $P_{\mathrm{u,n}}$ are the peak amplitude of the *n*-th harmonic and the *n*-th ultraharmonic frequency components, respectively, and $\bar{A}_{\mathrm{n}}$ is the averaged amplitude within the bandwidth of 75 kHz between the *n*-th harmonic and the *n*-th ultraharmonic frequencies. The 3rd to 6th harmonic/ultrahamonic frequencies were used. The frequency spectrum and the CDs were obtained for every burst. The CD was normalized by the electrical noise power that was obtained using the same processing pipeline but with electrical noise data. The electrical noise data were acquired without FUS transmission. The normalized CD was then converted to the logarithmic scale (i.e. dB) and presented in Figure 5A. The CCD was obtained by summing the normalized CD acquired after the microbubble flush and converting it to the logarithmic scale. We did not subtract the baseline from the CD or CCD due to high fluctuations in the baseline cavitation signals.


[1] Pouliopoulos AN, Kwon N, Jensen G, et al. Safety evaluation of a clinical focused ultrasound system for neuronavigation guided blood-brain barrier opening in non-human primates. Sci Rep. 2021; 11: 15043.

[2] Bae S, Liu K, Pouliopoulos AN, Ji R, Konofagou EE. Real-time passive acoustic mapping with enhanced spatial resolution in neuronavigation-guided focused ultrasound for blood-brain barrier opening. IEEE Trans Biomed Eng. 2023; 70: 2874–85.

### Supplementary Discussion

##### Pressure Estimation Errors

Although we aimed to deliver the same acoustic pressure (200 kPa) at the brain target, the variable BBBO volume across subjects indicated the pressure estimation errors in our pre-treatment simulation in the treatment planning step. The pressure estimation errors can be attributed to two main factors 1) CT-MR registration errors in the pre-treatment simulations and 2) transducer positioning errors during the treatment.

The predominant cause of the significant error in pressure estimation for subject 1 (i.e., 219% higher than the aimed pressure) was the CT-MR registration error—we noted the largest CT-MR registration error in subject 1's simulation. Additionally, the thicker skull of the subject 1 compared to other subjects might have also contributed to the large pressure estimation error. When excluding subject 1, the average pressure estimation error for the remaining subjects was 37.5%.

Aside from the CT-MR registration error, our analysis of 35 simulations per subject, which accounted for positioning errors of ±10 mm in distance and ±10° in angular deviation, revealed an average deviation in pressure estimates of 33%. Therefore, it is essential to maintain minimal transducer positioning error to ensure consistent BBBO outcomes.

### Supplementary Movie

**Movie S1.**

Trajectory of the focused ultrasound beam (blue) for blood–brain barrier opening treatment and the contrast-enhanced volume (green) after the treatment.
